# Effects of allocation concealment and blinding in trials addressing treatments for COVID-19: A methods study

**DOI:** 10.1101/2022.08.03.22278348

**Authors:** Dena Zeraatkar, Tyler Pitre, Juan Pablo Diaz-Martinez, Derek Chu, Bram Rochwerg, Francois Lamontagne, Elena Kum, Anila Qasim, Jessica J Bartoszko, Romina Brignardello-Petersen

**Author notes:** **Disclaimers:** None. **Data:** Available at https://osf.io/yqvus/. **Authors’ Contributions:** DZ conceived the study. DZ, TSP, EK, AQ, and JJB helped collect data. DZ, JPDM, and TSP performed analyses. DZ wrote the first draft of the manuscript. All authors reviewed and approved the final version of the manuscript.

## Abstract

**Objective:** Assess the impact of allocation concealment and blinding on the results of trials addressing COVID-19 therapeutics.

**Data sources:** World Health Organization (WHO) COVID-19 database and the Living Overview of the Evidence (L-OVE) COVID-19 platform by the Epistemonikos Foundation (up to February 4^th^ 2022)

**Methods:** We included trials that compared drug treatments, antiviral antibodies and cellular therapies with placebo or standard care. For the five most commonly reported outcomes, if sufficient data were available, we performed random-effects meta-regression comparing the results of trials with and without allocation concealment and trials in which both healthcare providers and patients were blinded with trials in which healthcare providers and/or patients were aware of the intervention. A ratio of odds ratios (ROR) > 1 or a difference in mean difference (DMD) > 0 indicates that trials without allocation concealment or open-label trials produced larger effects than trials with allocation concealment or blinded trials.

**Results:** As of February 4^th^ 2022, we have identified 488 trials addressing COVID-19 drug treatments and antiviral antibodies and cellular therapies. Of these, 436 trials reported on one or more of our outcomes of interest and were included in our analyses.

We found that trials without allocation concealment probably overestimate mortality (ROR 1.14 [95% CI 0.92 to 1.41]), need for mechanical ventilation (ROR 1.26 [95% CI 0.97 to 1.64]), admission to hospital (ROR 1.93 [95% CI 0.83 to 4.48]), duration of hospitalization (DMD 1.94 [95% CI 0.86 to 3.02]), and duration of mechanical ventilation (DMD 2.64 [95% CI −0.90 to 6.18]), but results were imprecise.

We did not find compelling evidence that double-blind and open-label trials produce consistently different results for mortality (ROR 1.00 [95% CI 0.87 to 1.15]), need for mechanical ventilation (ROR 1.03 [95% CI 0.84 to 1.26]), and duration of hospitalization (DMD 0.47 days [95% CI −0.38 to 1.32]). We found that open-label trials may overestimate the beneficial effects of interventions for hospitalizations (ROR 1.87 [95% CI 0.95 to 3.67] and duration of mechanical ventilation (DMD 1.02 days [95% CI −1.30 to 3.35]), but results were imprecise.

**Conclusion:** We found compelling evidence that, compared to trials with allocation concealment, trials without allocation concealment may overestimate the beneficial effects of treatments. We did not find evidence that trials without blinding addressing COVID-19 interventions produce consistently different results from trials with blinding. Our results suggest that consideration of blinding status may not be sufficient to judge risk of bias due to imbalances in co-interventions. Evidence users may consider evidence of differences in co-interventions between trial arms when judging the trustworthiness of open-label trials. We suggest, however, evidence users to remain skeptical of trials without allocation concealment.

**What’s new?:** *key findings:* Trials without blinding did not produce consistently different results from trials with blinding.

*Additional information:* Previous studies have had conflicting results with regards to the effects of blinding on trial results. Our study supports the assertion that results from blinded trials may not differ significantly from unblinded ones.

*Implications:* Our study suggest that risk of bias assessment of blinding needs to be more nuanced and that lack of blinding may not be a definite indication of risk of bias.

## Introduction

Randomized trials represent the optimal design for assessing the effectiveness of interventions. Randomization aims to balance prognostic factors between arms such that any observed differences between trial arms can be attributed to the intervention under study.

Blinding—the concealment of the arm to which participants have been randomized from one or more individuals involved in a trial—has long been considered important to prevent bias in randomized trials (1, 2). Blinding of participants and healthcare providers reduces opportunity for differences in care (co-interventions) whereas blinding of outcome assessors and adjudicators reduces differences in the measurement and adjudication of outcomes between trial arms (3, 4). Preconceived notions about the efficacy and safety of interventions, even subconsciously, may theoretically impact decisions about co-interventions and outcome expectancy. Though there are clear advantages, blinding requires significant resources, increases the operational complexity of trials, and may not always be feasible.

Allocation concealment—ensuring that trial personnel enrolling participants are unaware of the sequence of assignments—is also considered important to prevent bias in trials and eliminates the opportunity for investigators to assign participants to trial arms based on characteristics that may affect their outcomes.

Meta-epidemiological studies (studies that analyze the results of previous studies to address how methodological characteristics of the studies influence their results) have yielded inconsistent findings with regards to the effects of blinding on trial results, suggesting that the effects of blinding may depend on other contextual factors such as the disease and intervention under investigation, setting, and outcome (5–10). Further, while earlier work suggests that trials without allocation concealment may overestimate trial results, there is limited recent empirical data on the effects of allocation concealment on trial results and there is no data specifically addressing COVID-19 trials (11, 12)(10).

Trials addressing COVID-19 treatments were designed rapidly and attempted to answer questions efficiently. Seminal COVID-19 trials, for example, representing massive international collaborations, such as RECOVERY (13–16) and SOLIDARITY (17, 18), use open-label (unblinded) designs. Clarification of the circumstances in which blinding and allocation concealment are important, and an empirical assessment of direction and degree of bias when they are not implemented, has important implications for the interpretation of trial results and for the design of future trials—particularly for future health emergencies in which trials may be designed rapidly with limited funding. In this study, we assess whether blinded trials and trials with allocation concealment addressing COVID-19 treatments produce results that are different than open-label trials and trials without allocation concealment.

## Methods

We registered a protocol for this study at the Open Science Framework (OSF) (osf.io/yqvus/) (Supplement 1). Figure 1 presents a general overview of the methods.

**Figure 1:**
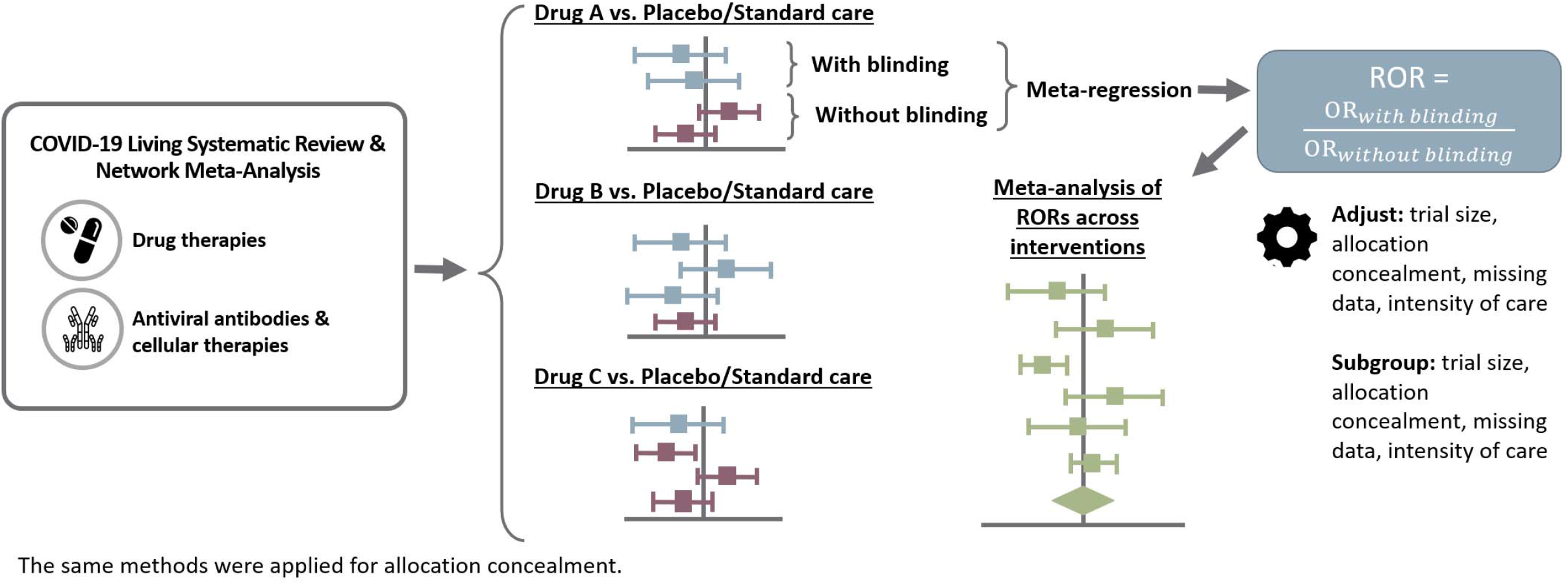
Overview of methods.

### Search Strategy

We used the search strategy of a linked living systematic review and living network meta-analysis (SRNMA) addressing therapeutics for COVID-19. Briefly, as part of this living review, we perform daily searches of the World Health Organization (WHO) COVID-19 database—a comprehensive multilingual source of global published and preprint literature on COVID-19 (https://search.bvsalud.org/global-literature-on-novel-coronavirus-2019-ncov/). Prior to its merge with the WHO COVID-19 database on 9 October 2020, we searched the US Centers for Disease Control and Prevention (CDC) COVID-19 Research Articles Downloadable Database. A validated machine learning model facilitates the efficient identification of randomized trials (19).

The search is supplemented by ongoing surveillance of living evidence retrieval services: the Living Overview of the Evidence (L-OVE) COVID-19 platform by the Epistemonikos Foundation (https://app.iloveevidence.com/loves/5e6fdb9669c00e4ac072701d) and the Systematic and Living Map on COVID-19 Evidence by the Norwegian Institute of Public Health (https://www.fhi.no/en/qk/systematic-reviews-hta/map/). Supplement 2 presents additional details of our search strategy.

### Study Selection

As part of the living SRNMA, pairs of reviewers, following calibration exercises, work independently and in duplicate to screen titles and abstracts of search records and subsequently the full texts of records deemed potentially eligible at the title and abstract screening stage. Reviewers resolve discrepancies by discussion or, when necessary, by adjudication with a third-party reviewer.

We include preprint and peer-reviewed reports of trials that randomize patients with suspected, probable, or confirmed COVID-19 to drug treatments, antiviral antibodies and cellular therapies, placebo, or standard care or trials that randomize healthy participants exposed or unexposed to COVID-19 to prophylactic drugs, standard care, or placebo. We do not apply any restrictions based on severity of illness, setting, or language of publication. We exclude trials that report on nutritional interventions, traditional Chinese herbal medicines without standardization in formulations and dosing across batches, and non-pharmacologic treatments.

For this study, we include all eligible trial reports of drug treatments and antiviral antibodies and cellular therapies up to February 4^th^, 2022. Because we anticipate that the impact of blinding may be different in trials addressing prophylaxis compared to trials addressing treatments, we exclude trials addressing prophylaxis (20).

### Data Collection

As part of the living SRNMA, for each eligible trial, pairs of reviewers, following training and calibration exercises, work independently to extract trial characteristics (country, publication status, funding), patient characteristics (inpatient vs. outpatient), methods (allocation concealment, blinding, missing outcome data), and results (number of participants and events in each arm for dichotomous outcomes and number of participants, mean of median number of participants, and standard deviation, standard error, or interquartile range for each arm for continuous outcomes) using a standardized, pilot tested data extraction form.

To assess risk of bias, pairs of reviewers, following training and calibration exercises, independently use a revision of the Cochrane tool for assessing risk of bias in randomized trials (RoB 2.0), presented in Supplement 3 (14). Reviewers resolve discrepancies by discussion and, when necessary, by adjudication with a third party.

For this study, we focus on trials that report on one or more of our outcomes of interest: mortality, mechanical ventilation, hospital admission, duration of hospitalization, and duration of mechanical ventilation. These outcomes represent the five most commonly reported outcomes among included trials.

We considered a trial to have adequate allocation concealment if the trial report described central randomization either via a computer or telephone system, pharmacy-controlled randomization, or sequentially-numbered opaque sealed envelopes (11).

We considered a trial blinded if patients and healthcare providers are described as being unaware of the intervention to which patients were assigned and described adequate blinding methods (i.e., matching placebo). We considered a trial open-label if patients and healthcare providers were described as being aware of the intervention to which patients were assigned. We planned to investigate the effects of blinding of patients and healthcare providers individually on results. We found, however, only a handful of trials that only blinded patients or only blinded healthcare providers and hence only compare trials in which both patients and healthcare providers and open-label trials. Hence, we compared trials in which both patients and healthcare providers were blinded with trials in which neither patients and healthcare providers were blinded or only one party was blinded.

For trial reports that did not clearly describe blinding or allocation concealment status, we applied validated guidance to determine the most likely blinding and allocation concealment status (21). For example, we assumed that both patients and healthcare providers were blinded when the trial described a placebo or a dummy medication or procedure. We assumed that trials described as ‘single-blind’ only blinded patients and trials described as ‘double-blind’ or ‘triple-blind’ blinded patients and healthcare providers. We assumed that trials that omitted any description of allocation concealment and blinding did not implement allocation concealment and blinding with one exception. We assumed that trials in which healthcare providers were blinded implemented allocation concealment because it is unlikely that healthcare providers could be adequately blinded without allocation concealment.

### Data Synthesis

We report categorical trial and patient characteristics as proportions and percentages and continuous characteristics as medians and interquartile ranges (IQR).

To assess the effects of allocation concealment and blinding on trial results, for each outcome and comparison (i.e., unique treatment vs. placebo/standard care) that included at least one trial with and one trial without allocation concealment or one trial in which patients and healthcare providers were blinded and one trial in which patients and healthcare providers were not blinded, we used random-effects meta-regressions with the restricted maximum likelihood (REML) estimator to estimate the ratios of odds ratios (RORs) or differences in mean differences (DMDs) corresponding to the difference in results between trials with blinding or allocation concealment compared to trials without double-blinding or allocation concealment. Subsequently, we combined RORs or DMDs using random-effects meta-analysis, with the REML heterogeneity estimator (22).

The effects of allocation concealment and blinding were coded such that an ROR greater than 1 indicates that, on average, trials without allocation concealment or open-label trials overestimate the beneficial effects of treatments compared to trials with allocation concealment or blinded trials. Similarly, DMD greater than 0 indicates that, on average, trials without allocation concealment or open-label trials overestimate the beneficial effects of treatments compared to trials with allocation concealment or blinded trials (Box 1).

#### Box 1

How to interpret the results

Odds ratio (OR) = Odds _intervention_/Odds _placebo/standard care_

ROR = OR _trials with allocation concealment or blinding_/ OR _trials without allocation concealment or blinding_

ROR > 1: trials without allocation concealment or blinding overestimate beneficial effects

ROR < 1: trials with allocation concealment or trials with blinding overestimate beneficial effects

ROR = 1: trials without allocation concealment or trials without blinding produce similar estimates to double-blind trials or trials with allocation concealment

Mean difference (MD) = Mean _intervention_ – Mean _placebo/standard care_

DMD = MD _trials with allocation concealment or blinding_ – MD _trials without allocation concealment or blinding_

DMD > 0: trials without allocation concealment or blinding overestimate beneficial effects

DMD < 0: trials with allocation concealment or blinding overestimate beneficial effects

DMD = 0: trials without allocation concealment or blinding produce similar estimates to trials with allocation concealment or blinding1

We hypothesized that the effects of allocation concealment and blinding may be influenced by blinding status for allocation concealment and allocation concealment status for blinding, number of patients randomized, risk of bias due to missing outcome data, and setting (inpatient versus outpatient). We conducted secondary analyses in which we adjusted meta-regression models for these variables.

To test whether the effects of blinding and allocation concealment may differ based on trial size, allocation concealment status for blinding and blinding status for allocation concealment, missing outcome data, and setting, we performed subgroup analyses comparing the effects of blinding and allocation concealment in trials with fewer than the median and equal to or more than the median number of participants, trials with allocation concealment versus without allocation concealment for blinding and trials in which patients and healthcare providers were blinded versus trials in which patients or healthcare providers were not blinded for allocation concealment, trials at low versus high risk of bias for missing outcome data, and inpatient versus outpatient trials.

We considered an ROR of 1.10 or 0.90 or a DMD of 1 day to indicate minimally important effects of allocation concealment and blinding on trial results. Assuming a baseline risk of 130 per 1,000 people for mortality, an overestimation of the odds ratio for the comparison between a treatment and standard care/placebo by 10% would result in an important overestimation of the absolute treatment effect (33 more per 1,000 people)—an effect that exceeds the 2% threshold of important effect (23).

We conducted all analyses in R (version 4.03, R Foundation for Statistical Computing), using the *meta* and *metafor* packages. Open Science Framework (https://osf.io/yqvus/) presents the data and code to generate the results in this study.

## Results

### Trial Characteristics

As of February 4^th^ 2022, we identified 488 trials addressing COVID-19 drug treatments and antiviral antibodies and cellular therapies. Of these, 436 trials reported on one or more of our outcomes of interest. Supplement 4 presents additional details about the selection of eligible trials.

We identified 352 trials (122 comparisons), 193 trials (79 comparisons), 62 trials (40 comparisons), 171 trials (80 comparisons), and 49 trials (25 comparisons) that included a placebo/standard care arm and that were eligible to be included in the analyses for mortality, mechanical ventilation, hospitalization, duration of hospitalization, and duration of mechanical ventilation, respectively.

Table 1 presents characteristics of trials that reported on mortality and mechanical ventilation and Supplement 5 presents characteristics of trials that reported on other outcomes.

**Table 1:**
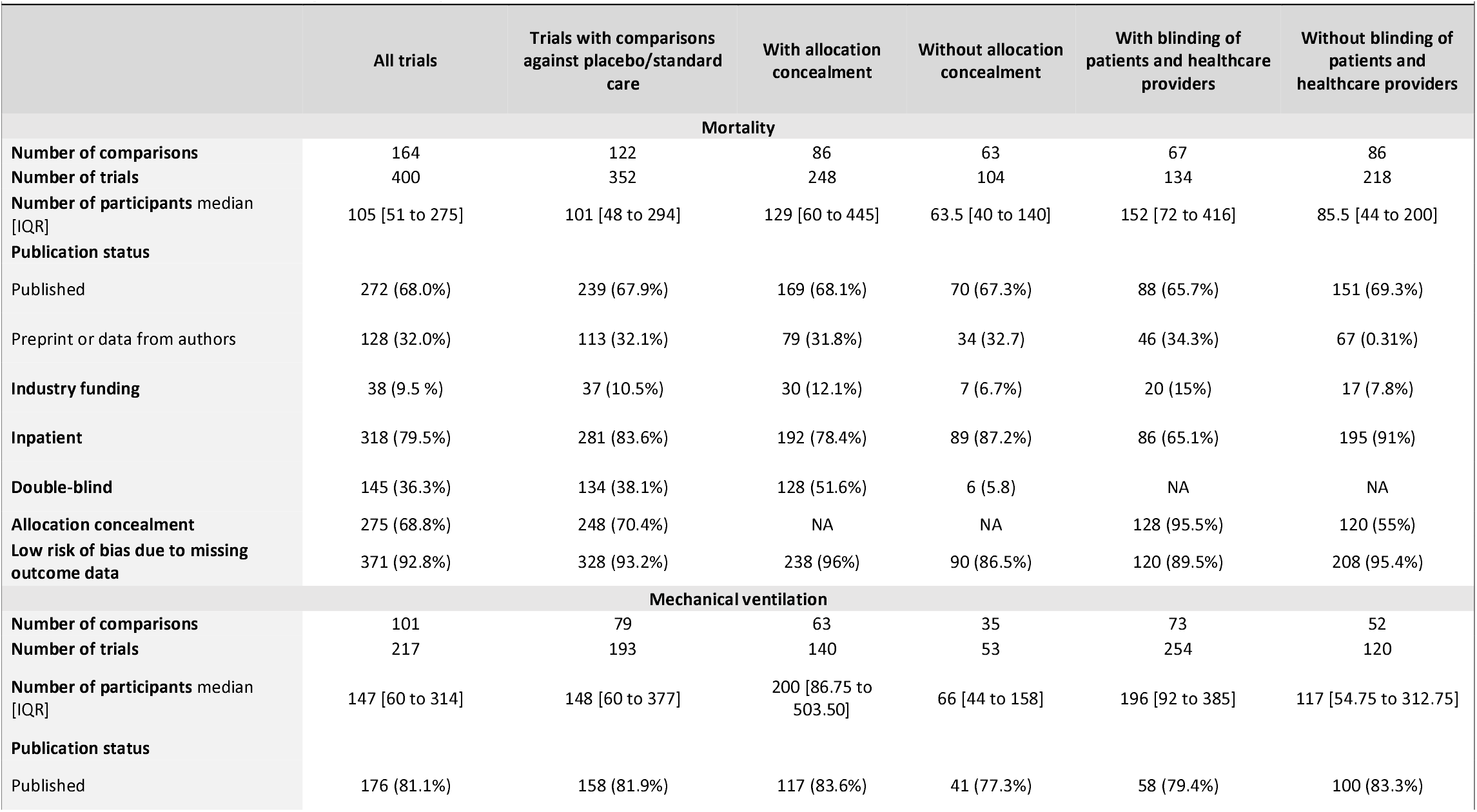
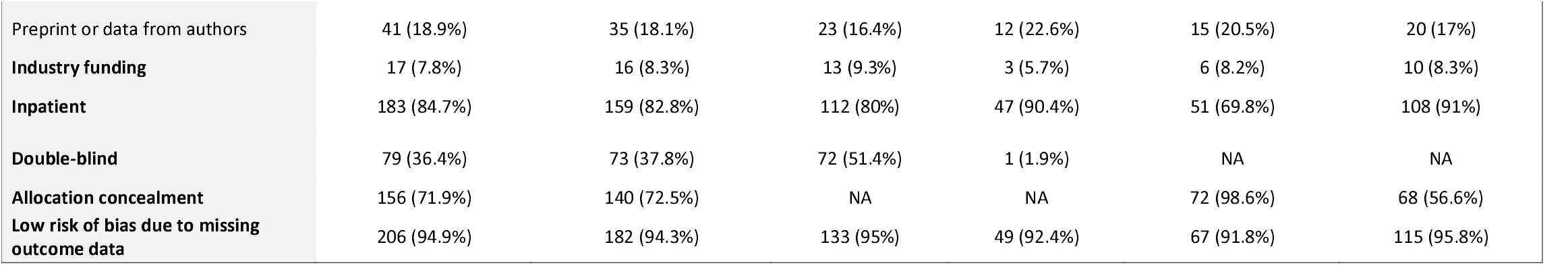
Characteristics of trails that report on mortality and mechanical ventilation.

We did not identify any important differences in the characteristics of trials that compared one or more active interventions with placebo or standard care and trials that compared different active interventions without a placebo or standard care arm.

Two thirds of these trials were published in peer-reviewed journals and the remainder were available as preprints or as part of systematic reviews and meta-analyses. Approximately one in ten trials were industry-funded or at high risk of bias for missing outcome data. While more than two-thirds of trials had allocation concealment, only a third blinded healthcare providers and patients.

We did not identify any important differences in the characteristics of trials that compared one or more active interventions with placebo or standard care and trials that compared different active interventions without a placebo or standard care arm.

Compared to trials without allocation concealment, trials with allocation concealment recruited more participants and were twice as likely to be industry-funded. Compared to trials without blinding, trials with blinding recruited more participants and were twice as likely to be industry-funded and slightly more likely to recruit outpatients. Blinding and allocation concealment status were correlated with most trials with blinding also having allocation concealment but only half of open-label trials having allocation concealment.

### Allocation Concealment

Compared to trials with allocation concealment, trials without allocation concealment probably overestimate improvements in mortality (ROR 1.14 [95% CI 0.92 to 1.41]; I^2^ = 0%), mechanical ventilation (ROR 1.26 [95% CI 0.97 to 1.64]; I^2^ = 0%), hospitalization (ROR 1.93 [95% CI 0.83 to 4.48]; I^2^ = 0%), duration of hospitalization (DMD 1.94 days [95% CI 0.86 to 3.02]; I^2^ = 17%), and duration of mechanical ventilation (DMD 2.64 days [95% CI −0.90 to 6.18]; I^2^ = 0%) (Figure 2). Results, however, were imprecise and do not exclude the possibility that allocation concealment may have no important effect.

**Figure 2:**
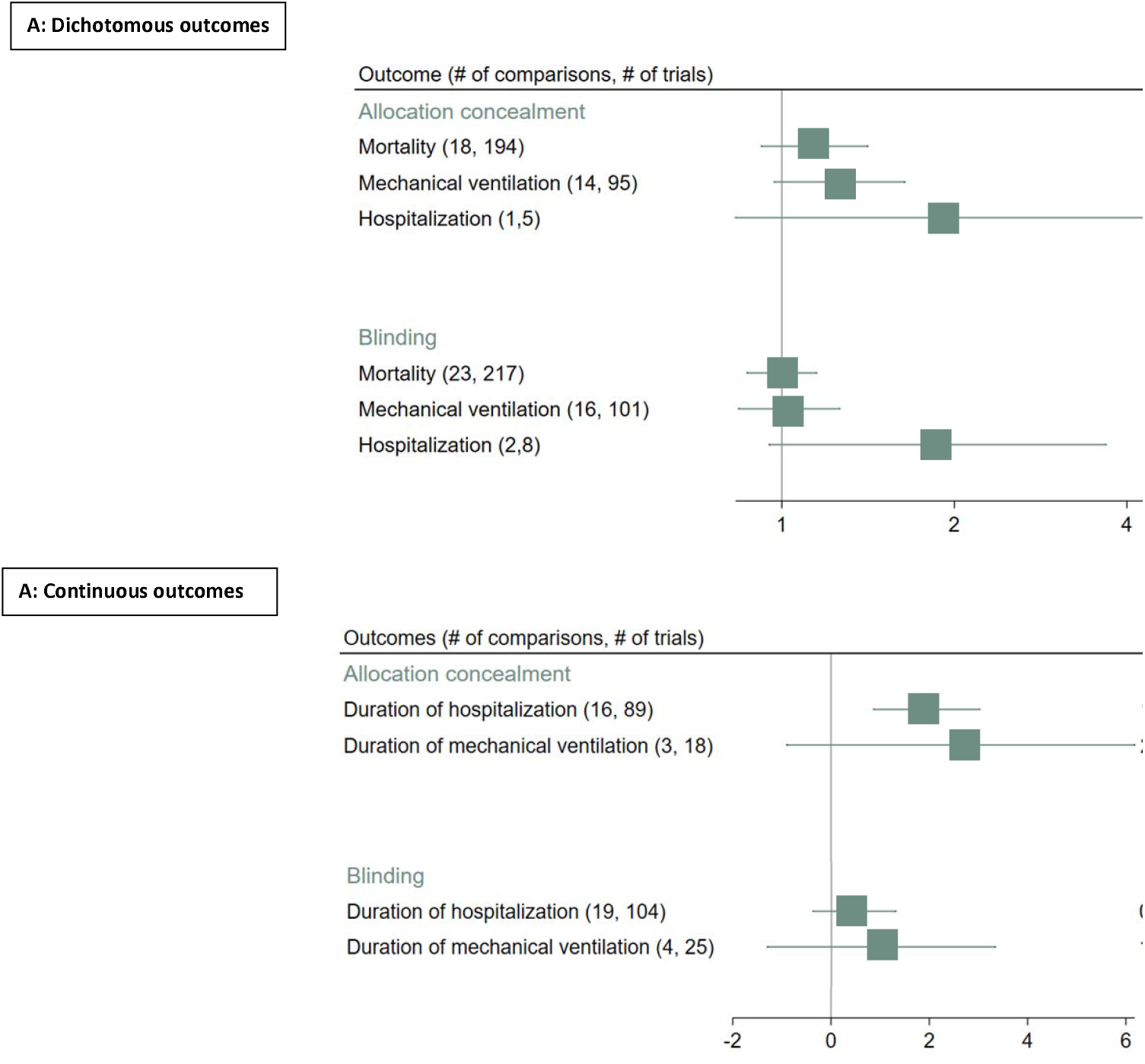
**Estimated ratio of odds ratios (RORs) and differences in mean differences (DMDs) comparing trials with and without blinding and trials with and without allocation concealment for dichotomous outcomes (A) and continuous outcomes (B); ROR 1 or DMD > 0: Trials without blinding or allocation concealment overestimate beneficial effects of interventions compared to trials with blinding or allocation concealment**

Figure 3 presents RORs comparing trials with and without allocation concealment across interventions for mortality. Supplement 6 presents figures for other outcomes.

**Figure 3:**
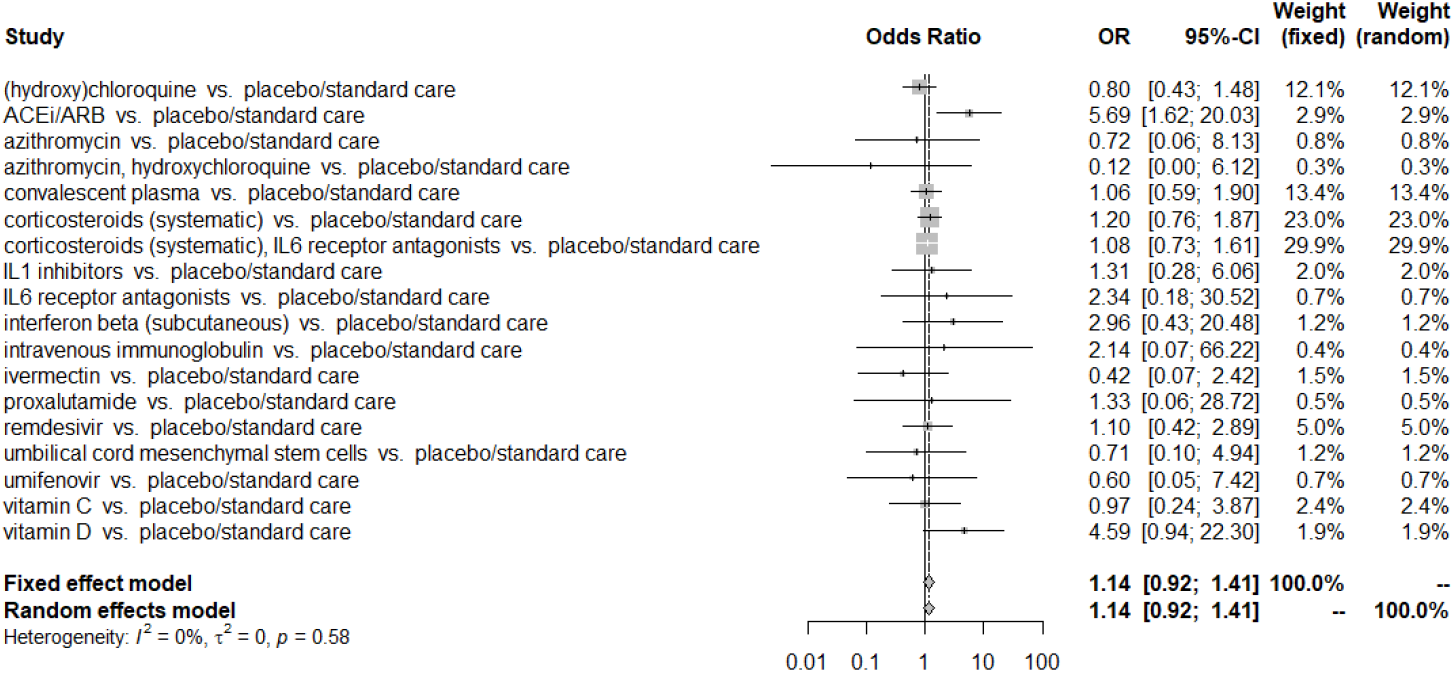
**Estimated ratios of odds ratios (RORs) comparing trials with and without allocation concealment reporting on COVID 19 treatments and mortality (ROR > 1: Trials without allocation concealment overestimate reduction in mortality c trials with allocation concealment)**

### Blinding

Compared to trials that blinded patients and healthcare providers, unblinded trials generated similar estimates for mortality (ROR 1.00 [95% CI 0.87 to 1.15]; I^2^ = 0%), need for mechanical ventilation (ROR 1.03 [95% CI 0.84 to 1.26]; I^2^ = 1%), and duration of hospitalization (DMD 0.47 days [95% CI −0.38 to 1.32]; I^2^ = 20%) (Figure 3). Unblinded trials, however, overestimate improvements for hospitalization (ROR 1.87 [95% CI 0.95 to 3.67]; I^2^ = 0%) and duration of mechanical ventilation (DMD 1.02 days [95% CI −1.30 to 3.35]; I^2^ = 71%).

Figure 4 presents RORs comparing trials with and without blinding across treatments for mortality. Supplement 10 presents figures for other outcomes.

**Figure 4:**
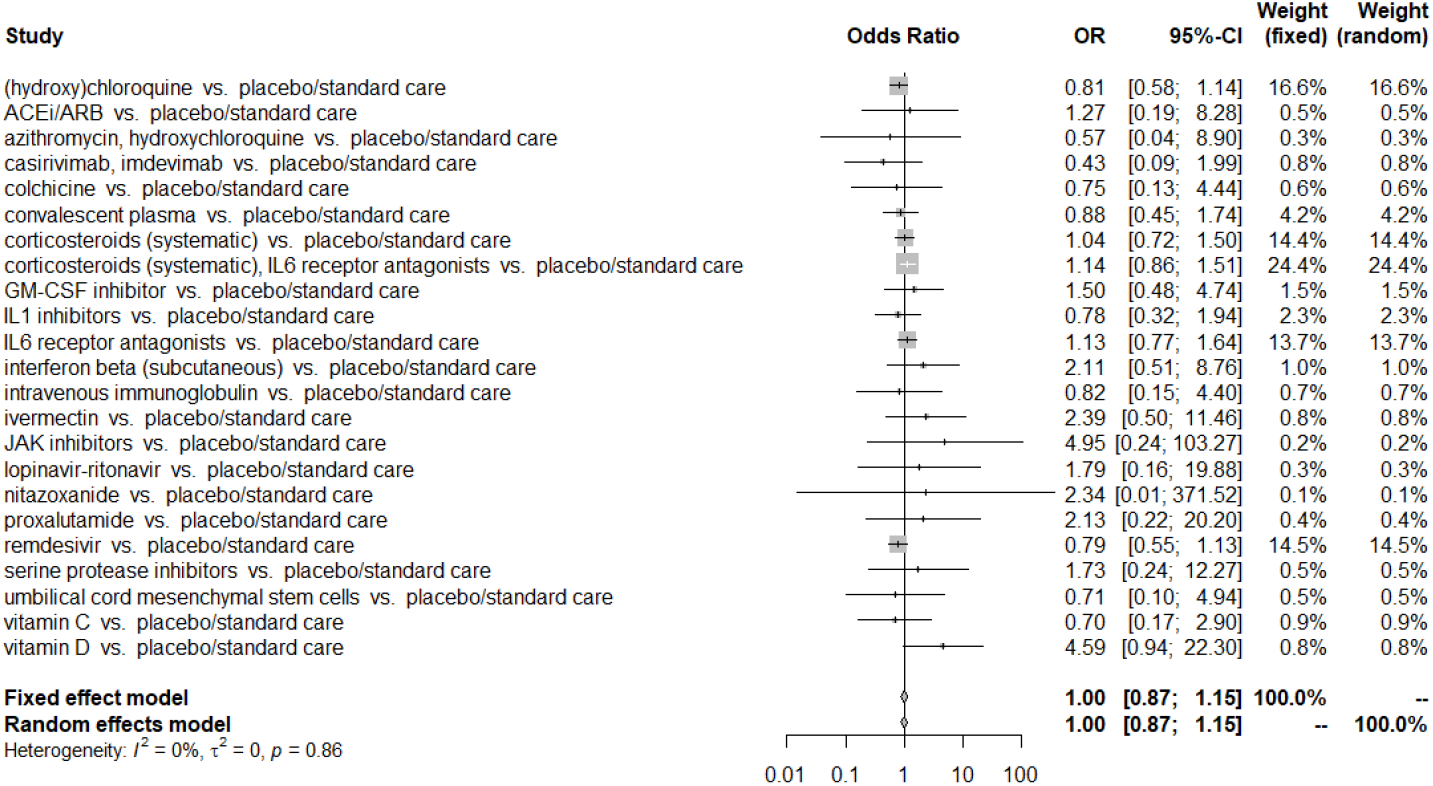
**Estimated ratios of odds ratios (RORs) comparing trials in which patients and healthcare providers were blinded and open-label trials reporting on COVID-19 treatments and mortality (ROR > 1: Trials without blinding trials overestimate reductio in mortality compared to trials with blinding)**

### Additional Analyses

Subgroup and sensitivity analyses, including adjusting for covariates and excluding studies not explicitly reporting allocation concealment, were consistent with the main findings (Supplements 7-9).

## Discussion

### Main Findings

Our study addresses the impact of allocation concealment and blinding on the results of trials addressing COVID-19 therapeutics.

Trials without allocation concealment, compared to trials with allocation concealment, probably overestimate the relative beneficial effects of interventions by an average of 14%, 26%, and 93% for mortality, mechanical ventilation, and hospitalizations, respectively, and on average by 2 and 3 days for duration of hospitalization and mechanical ventilation, respectively.

We did not find compelling evidence that trials with and without blinding produce consistently different results. Our findings, however, are subject to imprecision and do not exclude the possibility that open-label trials may produce biased results. Our results suggest that the effects of blinding may vary based on other trial characteristics (e.g., the intervention being compared and the type of outcome). For example, we found important heterogeneity in the estimated effects of blinding duration of mechanical ventilation.

### Relation to Previous Findings

These results are consistent with existing literature that suggests that the effects of blinding are modest—especially when outcomes are objective or outcomes assessors or adjudicators are blinded— and that trials without allocation concealment may produce biased results (10, 24).

Existing literature addresses the effects of blinding and allocation concealment in a variety of settings and so may not be directly applicable to COVID-19. A previous study addressing the effects of blinding on mortality in critical care RCTs found a modest effect of blinding (8).

### Strengths and Limitations

The strengths of this study include a large sample of trials addressing therapeutics for COVID-19, duplicate screening and extraction of data, and validated assumptions regarding blinding and allocation concealment status when they were incompletely reported in trials (21).

Our study also overcomes some of the limitations of the MetaBLIND study (5, 25). For example, almost half of the trials included in the MetaBLIND study came from systematic reviews of interventions for which we would not expect investigators to be able to administer co-interventions (i.e., reviews of interventions for informed consent and use of decision aids) (25). We restricted our analysis to comparisons of pharmacological, antiviral, and cellular treatments for COVID-19 for which imbalances in co-interventions are plausible.

Our results may be limited by confounding, whereby differences in characteristics of trials that are correlated with blinding and allocation concealment and that influence the magnitude of effect estimates distort the estimated effects of blinding and allocation concealment. We attempted to reduce the potential for confounding by adjusting for a set of a priori defined confounders.

To circumvent confounding, trial investigators may randomize trial participants to blind and open-label subtrials. Evidence from within trial comparisons of blinded and open-label subtrials suggest that open-label trials may overestimate effect estimates, but this evidence comes primarily from complementary/alternative medicine, predominantly acupuncture, and focuses on patient-reported outcomes (24). Hence, the observed results may be due to exaggeration of outcomes reported by patients with lack of blinding rather than due to possible imbalances in co-interventions.

Some of our analyses—particularly our subgroup analyses—were imprecise. Evidence users should be mindful that COVID-19 treatments have typically shown very modest effects and hence even small biases may affect results sufficiently to impact decision-making (23). Such modest effects may not easily be detected in analyses.

While we anticipate that trials without blinding and allocation concealment overestimate the beneficial effects of treatments, it is possible that the direction of bias caused by lack of blinding and allocation concealment may not be consistent. That is, in certain circumstances open-label trials may overestimate effects and in other circumstances they may underestimate effects. For example, in a situation in which investigators are not confident about the benefits of an intervention, they may administer more co-interventions to the patients assigned to the intervention arm. Conversely, in a situation in which investigators are more confident in the benefits of an intervention, they may be less attentive of patients assigned to the intervention arm. Differences in the effects of blinding and allocation concealment across interventions may have biased our results towards the null.

It is possible that trials that may be sensitive to biases due to imbalances in cointerventions implement blinding. Similarly, if trials with blinding produce more modest effect sizes compared to open-label trials, they may be more prone to publication bias and less likely to be included in this study, thus reducing observed differences between trials with and without blinding.

When blinding status and allocation concealment were not reported, we applied validated guidance to determine the most likely blinding status (21). It is, however, possible that we may have misclassified the status of some trials. We conducted sensitivity analyses excluding trials for which blinding status and allocation concealment were not clearly reported. Some of these sensitivity analyses produced imprecise results due to too few comparisons and trials.

Blinding may be compromised during a trial (e.g., due to obvious adverse events, poorly matched placebos, or large therapeutic effects), which may have attenuated the apparent difference between double-blind and open-label trials.

Finally, most trials to date have addressed interventions in severe-to-critical COVID-19, rather than mild-to-moderate COVID-19. While we attempted to test for differences in the effects of blinding and allocation concealment between inpatient versus outpatient trials, there were too few trials targeting outpatients. We did not include trials examining COVID-19 prophylaxis or vaccination or trials that compared active interventions without a placebo or standard care arm and so our results may not be applicable to these types of trials.

### Implications

Our findings have implications for the design of future COVID-19 trials and for evidence users who are interpreting and applying evidence from COVID-19 trials, such as systematic reviewers, guideline developers, and clinicians.

Our results support global efforts to streamline and simplify trials to be able to efficiently respond to health questions, even in the contexts of health emergencies (26–28). For decades, blinding has been considered fundamental to limit risk of bias in randomized trials (4, 5). Blinding, however, is costly, inconvenient, and sometimes not feasible. Partly due to these issues, seminal COVID-19 trials, for example, representing massive international collaborations, such as RECOVERY (13–16) and SOLIDARITY (17, 18), use open-label designs. Our results suggest that blinding or lack-there-of, does not preclude a trial from producing high certainty evidence.

For evidence users, our results suggest that judgements about the risk of bias caused by imbalances in co-interventions should include considerations beyond blinding status, such as whether co-interventions may influence outcomes, completeness in reporting of co-interventions, differences in co-interventions across trial arms and the possibility that differences in co-interventions may have influenced outcomes (Box 2)—considerations that are consistent with guidance from the Cochrane-endorsed risk of bias tool for assessing risk of bias due to deviations from intended intervention (29). For outcomes that are subjective, evidence users may also consider the blinding status of outcome assessors/adjudicators.

#### Box 2

Guidance for judging risk of bias related to imbalances in co-interventions in open-label trials

##### Is it possible that co-interventions may influence outcomes?

Co-interventions refer to interventions and supportive care, excluding the intervention being tested in the trial, that are administered to patients with the aim of improving health outcomes or preventing adverse health outcomes. Co-interventions are unlikely to influence the results of trials when there are no effective co-interventions that are accessible to the healthcare team and/or patients. Conversely, for conditions in which there are many other effective interventions that are accessible to the healthcare team and/or patients, co-interventions may influence outcomes.

For example, consider a trial for a novel gene therapy aimed at curing type I diabetes. Because there are no interventions that are known and accessible to the healthcare team and/or patients that could cure type I diabetes, evidence users need not be concerned about co-interventions. Conversely, in conditions like chronic pain or COVID-19 in which there are many possible co-interventions, including supportive therapies, that may affect outcomes, investigators should be concerned about the possibility that co-interventions may influence the results of open-label trials.

##### Are co-interventions across trial arms comprehensively reported?

If data on the administration of co-interventions that may influence outcomes across trial arms are comprehensively reported, evidence users can make judgements about how imbalances in co-interventions may have biased results. If such data are not reported, evidence users cannot make judgements about whether differences in co-interventions may have biased outcomes and may have to rely on blinding status to make judgements about potential for imbalances in co-interventions.

For example, an open-label trial addressing a novel therapy for COVID-19 should report the proportion of patients that received effective or potentially effective treatments or supportive therapies that may affect outcomes of interest. For COVID-19, such treatments include vaccinations, corticosteroids, IL-6 receptor blockers, JAK inhibitors, molnupiravir, nirmatrelvir/ritonavir, remdesivir, and supplemental oxygen or ventilation (30).

For evidence users to be able to make judgements about how co-interventions may have influenced outcomes, trials should at minimum report co-interventions that probably influence the outcomes of interest with moderate or high certainty (quality) beyond a certain threshold but the reporting of co-interventions with even low certainty would be optimal. The choice of threshold will depend on the anticipated magnitude of effect of the intervention under study. The smaller the anticipated magnitude of effect of the intervention, the smaller the threshold necessary for reporting co-interventions.

##### Are there important differences in co-interventions across trial arms? Is there a possibility that differences in co-interventions may have affected outcomes of interest?

Evidence users need to make judgements if there are differences in co-interventions that may influence the outcomes of interest across trial arms. To do this, evidence users need to consider both the magnitude of differences in co-interventions across trial arms and the magnitudes of effect of co-interventions on the outcomes of interest. Smaller imbalances in co-interventions are less likely to bias results compared to larger imbalances in co-interventions. Small imbalances in co-interventions with large effects are more likely to bias results compared to small imbalances in co-interventions with small effects.

To judge whether observed effects may be explained by differences in co-interventions across arms, evidence users should also consider the direction of the effect and the arm which received more potentially beneficial co-interventions. Differences in co-interventions are unlikely to have biased results in such a way to affect decision-making in a trial in which the intervention was found to be harmful and in which effective co-interventions were more commonly used in the intervention arm compared to the standard care arm.

To judge the extent to which imbalances in co-interventions may have biased trial results, evidence users may also consider “adjusting” trial results for imbalances in co-interventions to determine whether results change sufficiently to affect decision-making. Such an approach requires knowledge of the proportion of patients that received the co-intervention in either arm and the estimated effect of the co-intervention on the outcome of interest.

Consider a trial, for example, that finds a novel drug to reduce the relative risk for mortality by 30% compared to standard care (RR: 0.70, 95% CI 0.58 to 0.85). In this trial, however, more patients in the intervention arm compared to the control arm (20% vs. 10%) received a potentially effective co-intervention that reduces mortality by 20% (RR: 0.80). Evidence users can adjust the point estimate and the associated confidence intervals for the imbalance in co-interventions using the formula below. Adjustment for the imbalance in co-intervention yields a relative risk of 0.79 (95% CI 0.65 to 0.96). This represents the relative risk of the intervention compared to standard care adjusted for the imbalance in the co-intervention between arms.

##### Adjusted estimate

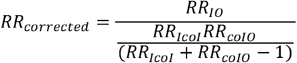

*RR*_*IO*_= intervention-outcome association

*RR*_*Icol*_= association between intervention and co-intervention (proportion in intervention arm exposed to co-intervention/proportion in comparator arm exposed to co-intervention)

*RR*_*coIo*_= co-intervention-outcome association

We refer readers to other sources that describe these methods in more detail (31). This approach, however, does not account for the uncertainty in the estimate of the effect of the co-intervention on the outcome of interest. Further, even with adjustment for imbalances in co-interventions, there may still be differences in co-interventions that are not accounted for or are not yet known to affect the outcome under study.

Our results suggest that allocation concealment should remain an important consideration in the design of trials and that evidence users should be skeptical of the results of trials in which allocation concealment was not implemented.

## Conclusion

We found compelling evidence that, compared to trials with allocation concealment, trials without allocation concealment probably overestimate the beneficial effects of treatments. We did not find compelling evidence that COVID-19 trials in which healthcare providers and patients are blinded produce consistently different results from open-label trials. Specific situations in which blinding may affect results, however, are unclear. We suggest evidence users to consider differences in co-interventions between trial arms when judging the trustworthiness of open-label trials but to remain skeptical of trials without allocation concealment.

## Data Availability

Can be found https://osf.io/yqvus/

## Acknowledgements

None

## Ethics statement

Not applicable, no human or animal subjects.

## Consent to participate and for publication

Not applicable

## Availability of data and material

Available at https://osf.io/yqvus/.

