## Supplementary material for "Effects of allocation concealment and blinding in trials addressing treatments for COVID-19: A methods study"

### Supplement 1 – Protocol

**Introduction**

Randomized trials represent the optimal design for assessing the effectiveness of interventions. Randomization ensures—more or less—that important prognostic factors are balanced between arms such that any observed differences between trial arms can be attributed to the intervention under study. For decades, blinding—the concealment of group allocation from one or more individuals involved in a trial—has been an important consideration in the assessment of risk of bias of randomized trials (1, 2). Blinding of participants and healthcare providers (hereon referred to as double-blinding) reduces opportunity in differences in care provided and sought whereas blinding of outcome assessors and adjudicators reduces differences in the measurement and adjudication of outcomes between trial arms (3, 4). Preconceived notions about the efficacy and safety of interventions, even subconsciously, may theoretically impact decisions about co-interventions and outcome expectancy. Blinding, however, requires significant resources, increases the operational complexity of trials, and may not always be feasible.

Allocation concealment—the concealment of the sequence to which participants will be randomized so that the group to which each subsequent participant will be randomized is unknown—is another important consideration in the assessment of the risk of bias of randomized trials. Theoretically, allocation concealment eliminates the opportunity for investigators to assign participants to trial arms based on characteristics that may affect their outcomes.

A meta-epidemiologic study of over a hundred meta-analyses did not find evidence that blinded and non- blinded trials produce systematically different results (5). The study, however, found important heterogeneity and produced imprecise estimates—suggesting that the effects of blinding may depend on other contextual factors such as the disease and intervention under investigation, setting, and outcome. Other meta-epidemiological studies have also produced inconsistent results (6, 7). Further, there is limited empirical data on the effects of allocation concealment.

Clarification of the circumstances in which blinding and allocation concealment are important, and an empirical assessment of direction and degree of bias when they are not implemented, has important implications for the interpretation of trial results and for the design of future trials.

Since March of 2022, we have performed a living systematic review and network meta-analysis (SRNMA) of treatments for COVID-19 (8, 9). To inform how we assessed the certainty of evidence, we considered whether open-label trials should be rated at high risk of bias either due to bias from imbalances in co-

interventions across arms or due to differences in measurement and adjudication of outcomes. Because our outcomes of interest are objective and not prone to expectancy effects, we did not consider measurement and adjudication of outcomes in open-label trials to be a potential source of bias. We were concerned, however, that even for objectives outcomes, differences in care and co-interventions may bias results—especially in the context of strong views from many healthcare providers on the efficacy or inefficacy of treatments being investigated (e.g., hydroxychloroquine, ivermectin). Further, we were concerned that trials without adequate allocation concealment may produce biased treatment effects by including patients with different characteristics in trial arms.

In this study, we plan to empirically assess whether double-blind trials and trials with allocation concealment addressing COVID-19 treatments produce results that are different than open-label trials and trials without allocation concealment for objective outcomes. We plan to investigate the effects of blinding of patients, blinding of healthcare providers, and blinding of both patients and healthcare providers on results.

### Methods

We will a protocol for this study at the Open Science Framework (OSF).

#### Search Strategy

Our study will use the search strategy of our living SRNMA. Briefly, we perform daily searches of the World Health Organization (WHO) COVID-19 database—a comprehensive multilingual source of global published and preprint literature on COVID-19 (https://search.bvsalud.org/global-literature-on-novel-coronavirus- 2019-ncov/). Prior to its merge with the WHO COVID-19 database on 9 October 2020, we searched the US Centers for Disease Control and Prevention (CDC) COVID-19 Research Articles Downloadable Database. A validated machine learning model facilitates the efficient identification of randomized trials (10).

Our search is supplemented by ongoing surveillance of living evidence retrieval services: the Living Overview of the Evidence (L-OVE) COVID-19 platform by the Epistemonikos Foundation (https://app.iloveevidence.com/loves/5e6fdb9669c00e4ac072701d) and the Systematic and Living Map on COVID-19 Evidence by the Norwegian Institute of Public Health ([https://www.fhi.no/en/qk/systematic-](https://www.fhi.no/en/qk/systematic-reviews-hta/map/) [reviews-hta/map/](https://www.fhi.no/en/qk/systematic-reviews-hta/map/)).

#### Study Selection

As part of the living SRNMA, pairs of reviewers, following calibration exercises, work independently and in duplicate to screen titles and abstracts of search records and subsequently the full texts of records

deemed potentially eligible at the title and abstract screening stage. Reviewers resolve discrepancies by discussion or, when necessary, by adjudication with a third-party reviewer.

We will include preprint and peer reviewed reports of trials that randomize patients with suspected, probable, or confirmed COVID-19 to drug treatments, antiviral antibodies and cellular therapies, placebo, or standard care or trials that randomize healthy participants exposed or unexposed to COVID-19 to prophylactic drugs, standard care, or placebo. We will not apply any restrictions based on severity of illness, setting, or language of publication. We will exclude trials that report on nutritional interventions, traditional Chinese herbal medicines without standardization in formulations and dosing across batches, and non-pharmacologic treatments.

For this study, we will include all eligible trial reports of drug treatments and antiviral antibodies and cellular therapies up to February 4^th^, 2022. Because we anticipate that the impact of blinding may be different in trials addressing prophylaxis compared to treatment (e.g., due to differences in preconceived notions about the effectiveness of prophylaxis and range of potentially effective and accessible co- interventions) we will exclude trials addressing prophylaxis (11).

#### Data Collection

As part of the living SRNMA, for each eligible trial, pairs of reviewers, following training and calibration exercises, independently will extract trial characteristics (country, publication status, funding), patient characteristics (inpatient vs. outpatient), methods (allocation concealment, blinding, missing outcome data), and results (number of participants and events in each arm for dichotomous outcomes and number of participants, mean of median number of participants, and standard deviation, standard error, or interquartile range for each arm for continuous outcomes) using a standardized, pilot tested data extraction form.

To assess risk of bias, pairs of reviewers, following training and calibration exercises, will independently use a revision of the Cochrane tool for assessing risk of bias in randomized trials (RoB 2.0) (14). Reviewers resolve discrepancies by discussion and, when necessary, by adjudication with a third party.

For this study, we will focus on trials report on one or more of our outcomes of interest: mortality, mechanical ventilation, hospitalization, duration of hospitalization, and duration of mechanical ventilation. These outcomes represent the five commonly reported patient-important outcomes.

We will consider a trial double-blinded if patients and healthcare providers are described as being unaware of the intervention to which patients were assigned and described adequate blinding methods

(i.e., matching placebo). We will consider a trial open label if patients and healthcare providers were aware of the intervention to which patients were assigned. We will consider a trial to have adequate allocation concealment if the trial report described central randomization either via a computer or telephone system, pharmacy-controlled randomization, or sequentially-numbered opaque sealed envelopes (12). For trial reports that do not clearly describe blinding or allocation concealment status, we will apply validated guidance to determine the most likely blinding and allocation concealment status (13).

#### Data Synthesis

We will report categorical trial and patient characteristics as proportions and percentages and continuous characteristics as medians and interquartile ranges (IQR).

We will restrict our analyses to trials that compare active interventions with placebo or standard care because preconceived notions about the effectiveness of alternative active interventions may be different, which could affect how co-interventions are administered.

To assess the effects of blinding and allocation concealment on trial results, for each outcome and comparison (i.e., treatment vs. placebo/standard care) that included at least one double-blind trial and one open label trial or one trial with and one trial without allocation concealment, we will use random- effects meta-regressions with the REML estimator to estimate the ratios of odds rations (RORs) or differences in mean differences (DMDs) corresponding to the difference between trials with blinding or allocation concealment compared to trials without blinding or allocation concealment. Subsequently, we will combine RORs or DMDs using random-effects meta-analysis, with the REML heterogeneity estimator (14).

The effects of blinding and allocation concealment will be coded such that an ROR greater than 1 indicates that, on average, open-label trials or trials without allocation concealment overestimate the beneficial effects of treatments compared to double-blind trials or trials with allocation concealment. Similarly, DMD greater than 0 indicates that, on average, open-label trials or trials without allocation concealment overestimate the beneficial effects of treatments compared to double-blind trials or trials with allocation concealment (Box 1).

**Box 1: How to interpret the results**

Odds ratio (OR) = Odds intervention/Odds placebo/standard care

ROR = OR double-blind or with allocation concealment/ OR open-label or without allocation concealment

ROR > 1: open-label trials or trials without allocation concealment overestimate beneficial effects

ROR < 1: double-blind trials or trials with allocation concealment overestimate beneficial effects

ROR = 1: open-label trials or trials without allocation concealment provide similar estimates to double- blind trials or trials with allocation concealment

Mean difference (MD) = Mean intervention – Mean placebo/standard care

DMD = MD double-blind or with allocation concealment – MD open-label or without allocation concealment

DMD > 0: open-label trials or trials without allocation concealment overestimate beneficial effects DMD < 0: double-blind trials or trials with allocation concealment overestimate beneficial effects DMD = 1: open-label trials or trials without allocation concealment provide similar estimates to double- blind trials or trials with allocation concealment

We hypothesize that the effects of blinding and allocation concealment may be influenced by allocation concealment status for blinding and blinding status for allocation concealment, trial size, missing outcome data, and setting (inpatient versus outpatient). We will conduct secondary analyses in which we adjust meta-regression models for these variables, first including each variable individually in the meta- regression models along with blinding or allocation concealment and subsequently including all variables simultaneously.

To test whether the effects of blinding and allocation concealment may differ based on trial size, allocation concealment status for blinding and blinding status for allocation concealment, missing outcome data, and setting, we will perform subgroup analyses comparing the effects of blinding and allocation concealment in trials with fewer than the median and equal to or more than the median number of participants, trials with allocation concealment versus without allocation concealment for blinding and trials with double-blinding versus without double-blinding for allocation concealment, trials at low versus high risk of bias for missing outcome data, and inpatient versus outpatient trials.

We will conduct all analyses in in R (version 4.03, R Foundation for Statistical Computing), using the *meta*

and *metafor* packages.

### Supplement 2 – Search strategy

**Search purpose**: Systematic search of the COVID-19 literature performed Monday through Friday for the WHO Database. Searches performed by Tomas Allen, Kavita Kothari, and Martha Knuth.

Use following commands to pull daily new entries:

• Entry_date:( [20210101 TO 20210120])

• Entry_date:( 20210105)

**Duplicates:** Duplicates are found in EndNote and Distillr using the Wichor method. Further screening is done by expert reviewers but some duplicates may still be in the database.

**Daily Search Strategy:**

| **Database** | **Search Strategy** |
| --- | --- |
| Medline  (Ovid)  1946- | (coronavir* OR corona virus* OR corona pandemic* OR betacoronavir* OR covid19 OR covid OR nCoV OR novel CoV OR CoV 2 OR CoV2 OR sarscov2 OR sars2 OR 2019nCoV OR wuhan virus*).mp. OR (sars AND cov).mp. OR ((wuhan OR hubei OR huanan) AND (severe acute respiratory OR pneumonia*) AND outbreak*).mp. OR Coronavirus Infections/ OR Coronavirus/ OR betacoronavirus/  Limits: 2020- |
| CAB Abstracts(Ovid)  1910- | (coronavir* OR corona virus* OR corona pandemic* OR betacoronavir* OR covid19 OR covid OR nCoV OR novel CoV OR CoV 2 OR CoV2 OR sarscov2 OR sars2 OR 2019nCoV OR wuhan virus*).mp. OR (sars AND cov).mp. OR ((wuhan OR hubei OR huanan) AND (severe acute respiratory OR pneumonia*) AND outbreak*).mp. OR exp Betacoronavirus/ |
| Global Health (Ovid)  1910- | (coronavir* OR corona virus* OR corona pandemic* OR betacoronavir* OR covid19 OR covid OR nCoV OR novel CoV OR CoV 2 OR CoV2 OR sarscov2 OR sars2 OR 2019nCoV OR wuhan virus*).mp. OR (sars AND cov).mp. OR ((wuhan OR hubei OR huanan) AND (severe acute respiratory OR pneumonia*) AND outbreak*).mp. OR exp Betacoronavirus/ |
| PsycInfo (Ovid)  1806- | (coronavir* OR corona virus* OR corona pandemic* OR betacoronavir* OR covid19 OR covid OR nCoV OR novel CoV OR CoV 2 OR CoV2 OR sarscov2 OR sars2 OR 2019nCoV OR wuhan virus*).mp. OR (sars AND cov).mp. OR ((wuhan OR hubei OR huanan) AND (severe acute respiratory OR pneumonia*) AND outbreak*).mp.  Limits: 2020- |
| Scopus  1960- | TITLE-ABS-KEY ( coronavir* OR "corona virus" OR "corona pandemic" OR betacoronavir* OR covid19 OR covid OR ncov OR "CoV 2" OR cov2 OR sarscov2 OR sars2 OR 2019ncov OR "novel CoV" OR "wuhan virus" ) OR TITLE-ABS-KEY(sars AND cov) OR ( TITLE-ABS-KEY ( wuhan OR hubei OR huanan ) AND TITLE-ABS-KEY ( "severe acute respiratory" OR pneumonia* ) AND TITLE-ABS-KEY ( outbreak* ) ) AND ( LIMIT-TO ( PUBYEAR , 2021 ) OR LIMIT-TO ( PUBYEAR , 2020 ) ) |
| Academic Search Complete (Ebsco) | TI,AB,SU( ( coronavir* OR "corona virus" OR "corona pandemic" OR betacoronavir* OR covid19 OR covid OR ncov OR "CoV 2" OR cov2 OR sarscov2 OR sars2 OR 2019ncov OR "novel CoV" OR "wuhan virus" ) OR (sars AND cov) OR ( ( wuhan OR hubei OR huanan ) AND ( "severe acute respiratory" OR pneumonia* ) AND ( outbreak* ) ) ) OR ( (MH "Coronavirus") OR (MH "Coronavirus Infections") ) Limits: Dec. 2019-, peer-reviewed |
| Africa Wide Information (Ebsco) | TI,AB,SU( ( coronavir* OR "corona virus" OR "corona pandemic" OR betacoronavir* OR covid19 OR covid OR ncov OR "CoV 2" OR cov2 OR sarscov2 OR sars2 OR 2019ncov OR "novel CoV" OR "wuhan virus" ) OR (sars AND cov) OR ( ( wuhan OR hubei OR huanan ) AND ( "severe acute respiratory" OR pneumonia* ) AND ( outbreak* ) ) ) Limits: 2019-, |
| CINAHL (Ebsco) | TI,AB,SU( ( coronavir* OR "corona virus" OR "corona pandemic" OR betacoronavir* OR covid19 OR covid OR ncov OR "CoV 2" OR cov2 OR sarscov2 OR sars2 OR 2019ncov OR "novel CoV" OR "wuhan virus" ) OR (sars AND cov) OR ( ( wuhan OR hubei OR huanan ) AND ( "severe acute respiratory" OR pneumonia* ) AND ( outbreak* ) ) ) OR ( (MH "Coronavirus") OR (MH "Coronavirus Infections") )  Limits: Dec. 2019-, peer-reviewed |
| ProQuest Central (Proquest)  1952- | TI,AB,SU( ( coronavir* OR "corona virus" OR "corona pandemic" OR betacoronavir* OR covid19 OR covid OR ncov OR "CoV 2" OR cov2 OR sarscov2 OR sars2 OR 2019ncov OR "novel CoV" OR "wuhan virus" ) OR (sars AND cov) OR ( ( wuhan OR hubei OR huanan ) AND ( "severe acute respiratory" OR pneumonia* ) AND ( outbreak* ) ) )  Limits: Dec. 2019-, peer-reviewed |
| China CDC MMWR | Covid OR cov2 OR coronavirus OR “sars cov” OR ncov |
| CDC Reports | Covid OR cov2 OR coronavirus OR “sars cov” OR ncov |
| bioRxiv  medRxiv  chemRxiv (preprints) | Covid OR cov2 OR coronavirus OR “sars cov” OR ncov |
| Embase (Ovid) | ncov OR (('coronavirus'/exp OR coronavirus) AND ('wuhan'/exp OR wuhan)) OR 'novel coronavirus' OR (('pneumonia'/exp OR pneumonia) AND wuhan:ti,ab) OR 'covid' OR 2019ncov OR 'sars-cov'/exp OR 'sars-cov' OR covid OR (('coronavirus'/exp OR coronavirus) AND novel) OR (('corona virus':ti,ab OR 'coronavirus':ti,ab) AND (outbreak:ti,ab OR epidemic*:ti,ab OR pandemdic*:ti,ab OR quaran*:ti,ab OR lockdown*:ti,ab OR syndemic*:ti,ab)) OR hcov OR 'sars virus'/exp OR 'sars virus' OR 'coronavirus disease 2019'/exp OR 'coronavirus disease 2019' OR 'novel coronavirus pneumonia' OR 'covid 19 virus' OR 'severe acute respiratory syndrome coronavirus 2'/exp OR 'severe acute respiratory syndrome coronavirus 2' OR 'coronavirinae'/exp OR 'coronavirinae' OR 'coronavirus infection'/exp OR 'coronavirus infection' OR 'covid19'/exp OR covid19 OR covid2019 OR 'corona pandemic' OR 'sarscov 2' OR 'sarscov-2' OR 'sars co v 2' OR 'social distancing'/exp OR 'social distancing' OR coivd OR 'flatten the curve' OR 'flattening the curve' OR pandoeconom* OR twindemic* OR 'sars voc' |
| Global Index Medicus | (nCov OR (coronavirus AND wuhan) OR "novel coronavirus" OR (pneumonia AND wuhan) OR covid OR 2019ncov OR "sars-cov " OR covid OR (coronavirus AND novel) OR (("corona virus" OR coronavirus ) AND ( ti:outbreak OR ti:epidemic* OR ti:pandemdic* OR ti:quaran* OR ti:syndem* OR hcov OR "sars virus")) OR "coronavirus disease 2019" OR " novel coronavirus pneumonia" OR "COVID 19 virus" OR "severe acute respiratory syndrome coronavirus 2" OR Coronavirinae OR "Coronavirus infection" OR covid19 OR covid2019 OR lockdown* OR "social distancing" OR “physical distancing” OR "corona pandemic" OR "sarscov 2" OR "sarscov-2" OR "sars co v 2" OR coivd OR "flatten the curve" OR "flattening the curve" OR "sars voc") |
| Web of Science | TI=coronavirus OR TI=covid OR TI=Covid19 OR TI=ncov OR TI=(SARS NEAR/3 COV) OR TI="novel coron*virus" OR TI=2019*ncoV OR TI=2019ncov OR TI=(CORON*VIRUS NEAR/3 (OUTBREAK OR pandemic OR 2019 OR new OR novel)) OR TI=coronavirinae OR TI=coronaviridae OR TI=betacoronavirus OR TI=Sars2 OR TI=COV2 OR TI=”corona pandemic” OR ((TI=wuhan OR TI=hubei OR TI=huanan) AND ( TI="severe acute respiratory" OR TI=pneumonia ) AND (TI=outbreak)) |
| PubMed Central | coronavirus[Title] OR "corona virus" [Title] OR "corona pandemic"[Title] OR coronavirinae[Title] OR coronaviridae[Title] OR betacoronavirus[Title] OR covid19[Title] OR covid[Title] OR nCoV[Title] OR "CoV 2"[Title] OR CoV2[Title] OR sars2[Title] OR sarscov2[Title] OR 2019nCoV[Title] OR "novel CoV"[Title] OR "wuhan virus"[Title] OR coronavirus[Abstract] OR "corona virus" [Abstract] OR "corona pandemic"[Abstract] OR coronavirinae[Abstract] OR coronaviridae[Abstract] OR betacoronavirus[Abstract] OR covid19[Abstract] OR covid[Abstract] OR nCoV[Abstract] OR "CoV 2"[Abstract] OR CoV2[Abstract] OR sars2[Abstract] OR sarscov2[Abstract] OR 2019nCoV[Abstract] OR "novel CoV"[Abstract] OR "wuhan virus"[Abstract] OR "COVID-19" [Supplementary Concept] OR "severe acute respiratory syndrome coronavirus 2" [Supplementary Concept] OR ((wuhan[Title] OR hubei[Title] OR huanan[Title]) OR (wuhan[Abstract] OR hubei[Abstract] OR huanan[Abstract]) AND ("severe acute respiratory"[Title] OR pneumonia[Title])) OR (("severe acute respiratory"[Abstract] OR pneumonia[Abstract]) AND (outbreak[Title]) OR outbreak[Abstract]) |
| Science Direct | COVID OR COVID19 OR 2019Ncov OR Ncov OR Coronavirus OR “corona virus” OR (SARS AND Cov) |
| Wiley Online | COVID-19 OR nCov OR 2019ncov OR (pneumonia AND wuhan) OR (sars AND cov) OR COVID OR Covid19 OR “corona virus” OR coronavirus OR COV2 OR SARS2 OR coronavirinae OR coronaviridae OR betacoronavirus OR "corona pandemic" OR ((wuhan OR hubei OR huanan) AND ( "severe acute respiratory" OR pneumonia ) AND (outbreak)) |

### Supplement 3 – Risk of Bias Guidance

| **Bias from the randomization process** | |
| --- | --- |
| Issues to consider:  Random sequence generation  Allocation concealment | |
| **Definitely low risk of bias** | Trials that assign participants to alternative interventions using a randomly generated sequence and maintain allocation concealment.  Examples of methods for developing a randomly generated allocation sequence include a random number generator, random number table, coin tossing, shuffling cards or envelopes, and throwing dice. If a trial is described as 'randomized' without any additional details related to how the allocation sequence was developed, we will assume that the allocation sequence was appropriately developed.  Examples of methods for maintaining allocation concealment include using central allocation via a computer or phone system, pharmacy-controlled allocation, opaque sealed envelopes, and sequentially numbered drug containers.  *Note that an explicit description of random sequence generation is not necessary for a rating of low risk of bias.* |
| **Probably low risk of bias** | Trials in which healthcare providers were blind to the intervention but which provide no information on allocation concealment and in which there are no major baseline imbalances.  *Note that an explicit description of random sequence generation is not necessary for a rating of probably low risk of bias.* |
| **Probably high risk of bias** | Trials in which healthcare providers were not blind to the intervention and which provide no information on allocation concealment.  Trials in which there are substantial baseline differences between trial arms that suggest a problem with the randomization process but there are no other limitations related to randomization. |
| **Definitely high risk of bias** | Trials in which allocation is by judgment of the clinician, by preference of the participant, by availability of the intervention, based on the results of a laboratory test, or other non-random rules (e.g., birthdate, etc.).  Trials in which investigators enrolling participants could possibly foresee the arm to which each subsequent patient would be randomized, such as allocation using an open allocation schedule (e.g. a list of random numbers), assignment envelopes used without appropriate safeguards (e.g. use of unsealed, non-opaque or not sequentially numbered envelopes), alternation between arms, case record number, or any other explicitly unconcealed procedure, rate as high risk. |
| **Bias due to deviations from the intended intervention** | |
| Issues to consider:  Blinding of healthcare providers/clinicians and participants  Imbalances in cointerventions or behaviors | |
| **Definitely low risk of bias** | Therapy trials in which healthcare providers are blind to the intervention administered and in which there are no significant differences in administered co-interventions.  Therapy trials that are described as double or triple blind.  Prophylaxis trials in which participants are blind to the intervention that they have been randomized.  Prophylaxis trials that are described as double or triple blind. |
| **Probably low risk of bias** |  |
| **Probably high risk of bias** | Therapy trials in which healthcare providers are not blind to the intervention administered.  Therapy trials in which healthcare providers are blind to the intervention administered but there are significant differences in administered co-interventions that suggests that blinding may have been compromised.  Therapy trials in which healthcare providers are described as being blind to the intervention but allocation concealment was inadequate.  Prophylaxis trials in which participants are not blind to the intervention that they have been randomized.  Prophylaxis trials in which participants are blind to the intervention to which they have been randomized but there are significant differences in social distancing and risk-taking behaviors that suggest that blinding may have been compromised.  Prophylaxis trials in which healthcare providers are not blind to the intervention and in which healthcare providers were very involved and counselled patients on social distancing, risk-taking behaviors, or testing for COVID-19. |
| **Definitely high risk of bias** | Therapy trials in which healthcare providers are not blind to the intervention and in which there are significant differences in administered co-interventions.  Prophylaxis trials in which participants are not blind to the intervention and in which there are significant differences in social distancing and risk-taking behaviors. |
| **Bias due to missing data** | |
| Issues to consider:  Missing outcome measures  Loss to follow-up | |
| **Definitely low risk of bias** | Trials in which missing outcome data (including outcome data that has been imputed) < 10%.  For in-patient trials, we will assume low risk of bias due to missing data unless otherwise specified. |
| **Probably low risk of bias** | Trials in which missing outcome data (including outcome data that has been imputed) is between 10% to 15% and missing outcome data is unlikely to be related to the true outcome and there is no imbalance in numbers of or reasons for missing data across intervention groups. |
| **Probably high risk of bias** | Trials in which missing outcome data (including outcome data that has been imputed) is between 10% to 15% and missing outcome data is likely to be related to the true outcome or there are imbalances in numbers of or reasons for missing data across intervention groups. |
| **Definitely high risk of bias** | Trials in which missing outcome data (including outcome data that has been imputed) > 15%. |
| **Bias due to measurement of the outcome** | |
| Issues to consider:  Blinding of outcome adjudicators  Objectivity of outcome  *Note that the judgments may differ across outcomes.* | |
| **Definitely low risk of bias** | Trials in which patients are blind to the intervention and in which outcomes are patient-reported.  Trials in which outcomes are measured by a third-party (investigator or clinician) and in which the third-party is blind to the intervention.  Trials in which the outcomes are objective (e.g., mortality, infection with COVID-19 confirmed by a positive RT-PCR swab, mechanical ventilation, admission to hospital, duration of hospital stay, ICU length of stay, ventilator free days, duration of mechanical ventilation, time to clinical improvement if clinical improvement is measured via objective criteria, viral clearance, time to viral clearance).  Trials that are described as double or triple blind. |
| **Probably low risk of bias** |  |
| **Probably high risk of bias** |  |
| **Definitely high risk of bias** | Trials in which patients are not blind and in which outcomes are patient-reported (e.g., time to symptom resolution).  Trials in which outcome adjudicators are not blind and the outcomes are not objective (e.g., adverse effects leading to discontinuation, transfusion-related acute lung injury, transfusion-associated circulatory overload, allergic reactions, infection with suspected/symptomatic COVID-19, venous thromboembolism, time to symptom resolution including fever, time to clinical improvement if the criteria for clinical improvement are not objective). |
| **Bias in selection of the reported results** | |
| Issues to consider:  Selective reporting of timepoints  Selective reporting of outcome measures  *Note that we are only interested in selective reporting for the outcomes for which we are extracting data.*  *Note that the judgments may differ across outcomes.* | |
| **Definitely low risk of bias** | Results for outcomes that were analyzed and reported according to a pre-specified statistical analysis plan or protocol (including the timepoint for the measurement of the outcome). |
| **Probably low risk of bias** | Results for outcomes that were analyzed and reported but that were not prespecified in a statistical analysis plan or protocol but the timepoint at which results are reported is consistent with the timepoint for other outcomes in the trial report or there is little reason to believe the outcome was selectively reported.  Please note that outcomes that were not prespecified in a protocol or statistical analysis plan and that are reported in the trial preprint or publication should be rated at probably low risk of bias unless there are other important reasons to suspect that results for those outcomes were selectively reported (e.g., results are presented at timepoints that don’t match the timepoints reported for other outcomes). |
| **Probably high risk of bias** | Results for outcomes that were analyzed and reported but that were not prespecified in a statistical analysis plan or protocol but the timepoint at which results are reported is not consistent with the timepoint for other outcomes in the trial report or there are other reasons to believe that the outcome is selectively reported. |
| **Definitely high risk of bias** | Results for outcomes that were analyzed and reported for which there are inconsistencies with the statistical analysis plan or protocol. These inconsistencies may include outcome measures of interest or the timepoints for the measurement of outcomes. |

### Supplement 4 – Study selection

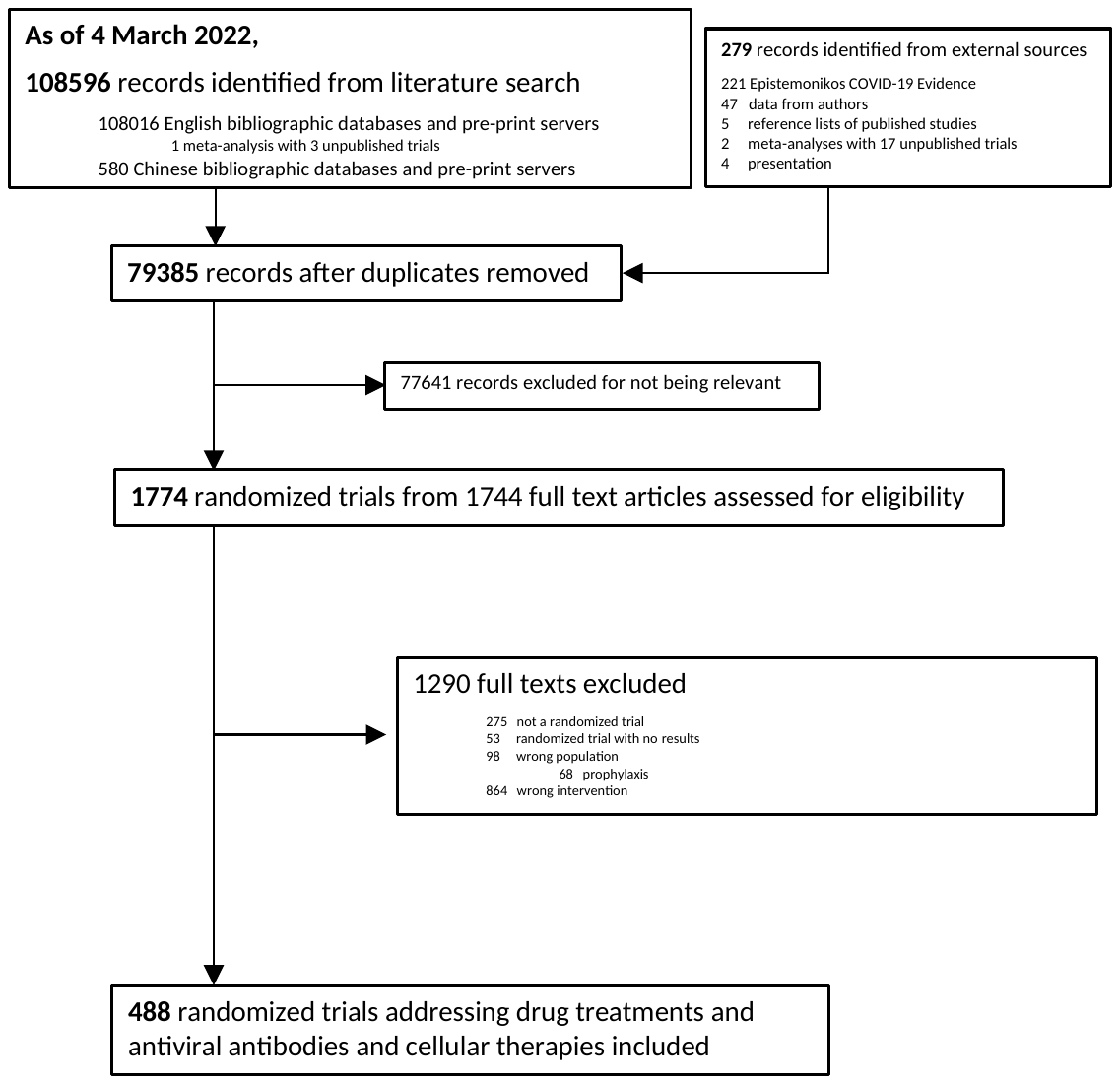

### Supplement 5 – Characteristics of trials that reported on hospitalization, duration of hospitalization, and duration of mechanical ventilation

| Characteristics of trials that reported on hospitalization, duration of hospitalization, and duration of mechanical ventilation | | | | | | |
| --- | --- | --- | --- | --- | --- | --- |
|  | **All trials** | **Trials with comparisons against placebo/standard care** | **Double-blind (patients and healthcare providers)** | **Open-label** | **With allocation concealment** | **Without allocation concealment** |
| Hospitalization | | | | | | |
| Number of comparisons | 49 | 40 | 31 | 11 | 34 | 8 |
| Number of trials | 72 | 62 | 47 | 15 | 53 | 9 |
| Number of participants median [IQR] | 214 [97.75 to 520] | 215 [100 to 558.25] | 300 [108.5 to 554.5] | 214 [80 to 575.5] | 307 [100 to 640] | 152 [60 to 214] |
| Publication status |  |  |  |  |  |  |
| Published | 46 (63.9%) | 41 (66.1%) | 31 (66%) | 10 (66.6%) | 35 (66%) | 6 (66.7%) |
| Preprint or data from authors | 26 (36.1%) | 21 (33.9%) | 16 (34%) | 5 (33.3%) | 18 (44%) | 3 (33.3%) |
| Industry funding | 8 (11.1%) | 7 (11.3%) | 5 (10.6%) | 2 (13.3%) | 6 (11.3%) | 1 (11.1%) |
| Inpatient | 3 (4.2%) | 3 (4.8%) | 1 (2.1%) | 2 (13.3%) | 1 (1.9%) | 1 (11.1%) |
| Allocation concealment | 61 (85%) | 53 (85.5%) | 46 (97.9%) | 7 (46.6%) | NA | NA |
| Patients and healthcare providers blinded | 51 (71%) | 47 (75.8%) | NA | NA | 46 (87%) | 1 (11.1%) |
| Low risk of bias due to missing outcome data | 65 (90.3%) | 58 (93.5%) | 43 (91.5%) | 15 (100%) | 49 (92.4%) | 9 (100%) |
| Duration of mechanical ventilation | | | | | | |
| Number of comparisons | 26 | 25 | 15 | 15 | 21 | 7 |
| Number of trials | 50 | 49 | 18 | 31 | 39 | 10 |
| Number of participants median [IQR] | 32 [14.50 to 74.75] | 33 [16 to 79] | 48 [22.25 to 89.75] | 32 [11 to 68.5] | 40 [20 to 100] | 14 [10 to 41.5] |
| Publication status |  |  |  |  |  |  |
| Published | 38 (76%) | 38 (77.5%) | 14 (78%) | 24 (77.4%) | 31 (79.5%) | 7 (70%) |
| Preprint or data from authors | 12 (24%) | 11 (22.4%) | 4 (22.2%) | 7 (22.6%) | 8 (20.5%) | 3 (30%) |
| Industry funding | 7 (14%) | 7 (14.3%) | 1 (5.5%) | 6 (19.3%) | 6 (15.4%) | 1 (10%) |
| Inpatient | 49 (98%) | 48 (98%) | 17 (94.4%) | 31 (100%) | 38 (97.4%) | 10 (100%) |
| Allocation concealment | 40 (80%) | 39 (79.6%) | NA | 21 (67.7%) | NA | NA |
| Patients and healthcare providers blinded | 18 (36%) | 18 (36.7%) | NA | NA | 18 (46.1%) | NA |
| Low risk of bias due to missing outcome data | 49 (98%) | 48 (97.9%) | 18 (100%) | 30 (96.8%) | 38 (97.4%) | 10 (100%) |
| Duration of hospitalization | | | | | | |
| Number of comparisons | 102 | 80 | 42 | 56 | 55 | 42 |
| Number of trials | 196 | 171 | 57 | 114 | 114 | 57 |
| Number of participants median [IQR] | 99 [50 to 238] | 93 [49.5 to 229.5] | 117 [56 to 314] | 81.50 [48.25 to 200] | 125.50 [60.25 to 408.25] | 65 [41 to 100] |
| Publication status |  |  |  |  |  |  |
| Published | 157 (80.1%) | 141 (82.4%) | 48 (84.2%) | 93 (81.6%) | 95 (83.3%) | 46 (81%) |
| Preprint or data from authors | 39 (19.9%) | 30 (17.5%) | 9 (15.8%) | 21 (18.4%) | 19 (16.7%) | 11 (19.3%) |
| Industry funding | 16 (8.2%) | 15 (8.8%) | 6 (10.5%) | 9 (7.9%) | 12 (10.5%) | 3 (5.3%) |
| Inpatient | 183 (94.3%) | 160 (94.5%) | 51 (91.1%) | 109 (96.4%) | 105 (93%) | 55 (98.2%) |
| Allocation concealment | 128 (65.3%) | 114 (66.7%) | NA | 57 (50%) | NA | NA |
| Patients and healthcare providers blinded | 62 (32.6%) | 57 (33.3%) | NA | NA | 57 (50%) | NA |
| Low risk of bias due to missing outcome data | 185 (94.4%) | 161 (94.1%) | 53 (93%) | 108 (94.7%) | 109 (95.6%) | 52 (91.2%) |

### Supplement 6 – Estimated ratios of odds ratios (RORs) and differences in mean differences (DMDs) comparing trials with and without with and without allocation concealment

#### Mechanical ventilation

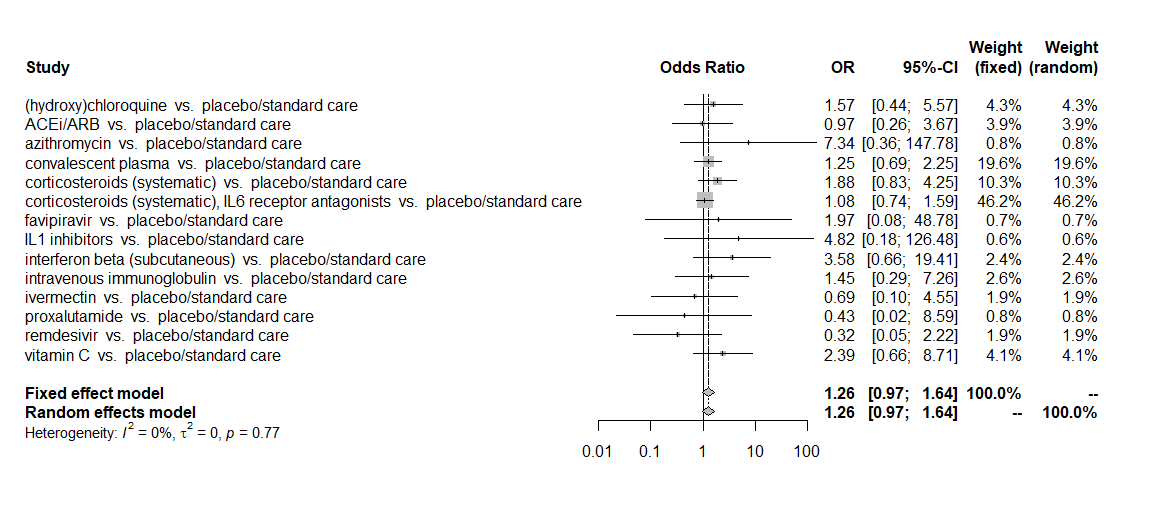

#### Hospitalization

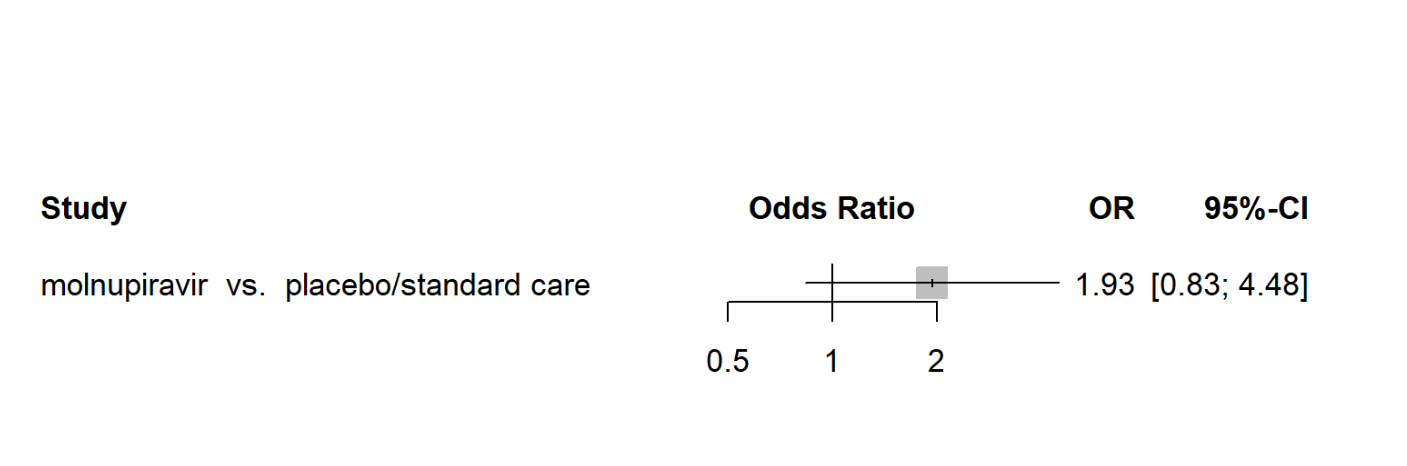

#### Duration of hospitalization

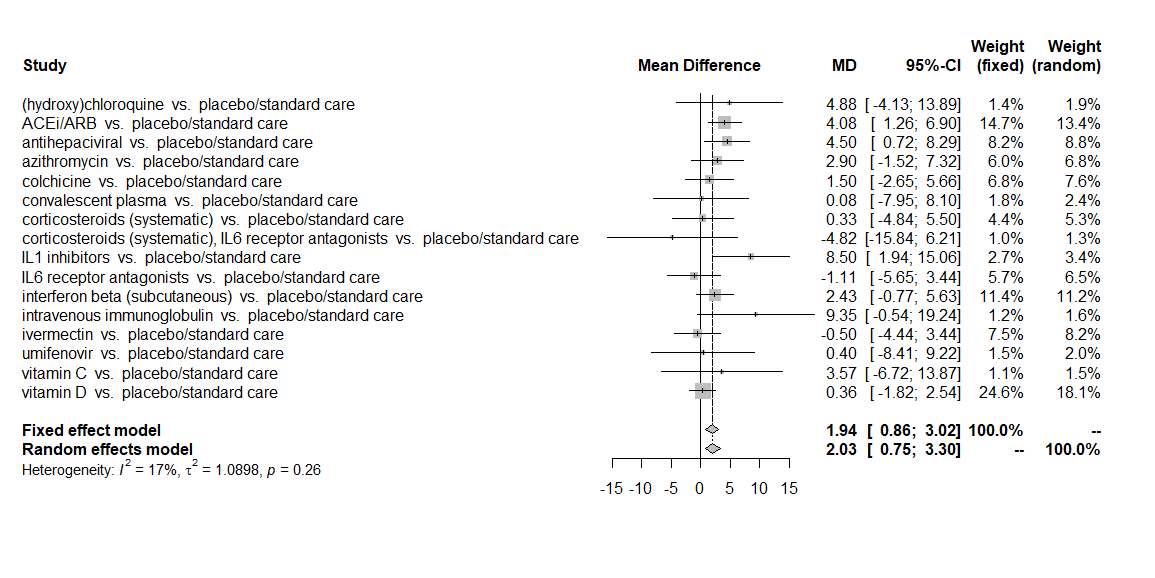

#### Duration of mechanical ventilation

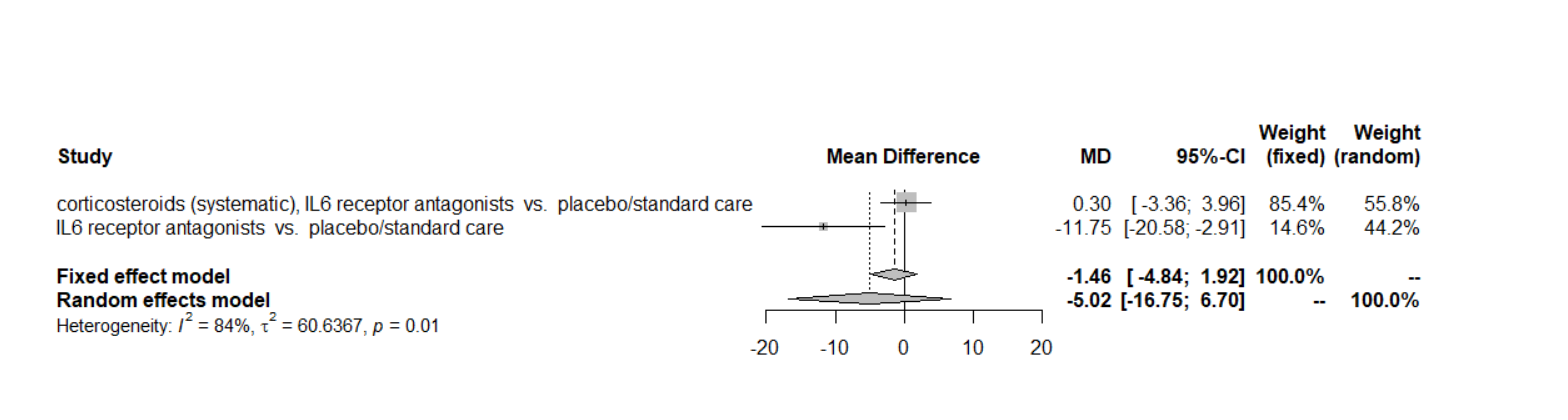

### Supplement 7 – The effects of allocation concealment and blinding on trial results across subgroups of trials

| **The effects of allocation concealment and blinding on trial results (ROR > 1 suggests that open-label trials or trials without allocation concealment overestimate beneficial effects of interventions compared to double-blind trials or trials with allocation concealment) across subgroups** | | | | | | | | | | | | | | | | |
| --- | --- | --- | --- | --- | --- | --- | --- | --- | --- | --- | --- | --- | --- | --- | --- | --- |
|  | **Mortality** | | | **Mechanical ventilation** | | | **Hospitalization** | | | **Duration of hospitalization** | | | **Duration of mechanical ventilation** | | | |
|  | **Comparisons (trials)** | **ROR (95% CI)** | **I2 (%)** | **Comparisons (trials)** | **ROR (95% CI)** | **I2 (%)** | **Comparisons (trials)** | **ROR (95% CI)** | **I2 (%)** | **Comparisons (trials)** | **DMD (95% CI)** | **I2 (%)** | | **Comparisons (trials)** | **DMD (95% CI)** | **I2 (%)** |
| **Allocation concealment** | | | | | | | | | | | | | | | | |
| All trials | 18 (194) | Random: 1.14 (0.92 to 1.41) Fixed: 1.14 (0.92 to 1.41) | 0 | 14 (95) | Random: 1.26 (0.97 to 1.64) Fixed: 1.26 (0.97 to 1.64) | 0 | 1 (5) | 1.93 (0.83 to 4.48) | 0 | 16 (89) | Random: 1.94 (0.86 to 3.02) Fixed: 2.03 (0.75 to 3.30) | 17 | | 3 (18) | Random: 2.64 (-0.90 to 6.18) Fixed: 2.64 (-0.90 to 6.18) | 0 |
| **Subgroups** |  |  |  |  |  |  |  |  |  |  |  |  | |  |  |  |
| Trial size |  |  |  |  |  |  |  |  |  |  |  |  | |  |  |  |
| < median | 12 (92) | Random: 1.24 (0.74 to 2.05) Fixed: 1.24 (0.74 to 2.05) | 0 | 8 (32) | Random: 1.65 (0.88 to 3.09) Fixed: 1.65 (0.86 to 3.14) | 9 | NA (NA) | NA | NA | 7 (28) | Random: 2.34 (0.24 to 4.43) Fixed: 2.34 (0.24 to 4.43) | 5 | | 1 (8) | 2.37 (-3.99 to 8.72) | 0 |
| ≥ median | 10 (75) | Random: 1.13 (0.87 to 1.48) Fixed: 1.13 (0.87 to 1.48) | 17 | 7 (38) | Random: 1.15 (0.88 to 1.51) Fixed: 1.15 (0.88 to 1.51) | 0 | NA (NA) | NA | NA | 5 (26) | Random: 3.31 (1.98 to 4.64) Fixed: 3.31 (1.97 to 4.65) | 1 | | 2 (4) | 5.66 (-0.29 to 11.6) | 0 |
| Blinding |  |  |  |  |  |  |  |  |  |  |  |  | |  |  |  |
| with blinding | 1 (21) | Random: 0.69 (0.31 to 1.56) Fixed: 0.69 (0.31 to 1.56) | 0 | NA (NA) | NA | NA | NA (NA) | NA | NA | 1 (4) | Fixed: -1.69 (-11.28 to 7.9) | 0 | | 0 (0) | NA | NA |
| without blinding | 13 (117) | Random: 1.15 (0.92 to 1.46) Fixed: 1.15 (0.92 to 1.46) | 0 | 12 (69) | Random: 1.22 (0.92 to 1.62) Fixed: 1.22 (0.92 to 1.62) | 0 | 1 (3) | 1.45 (0.45 to 4.69) | 0 | 11 (56) | Random: 2.82 (1.56 to 4.09) Fixed: 2.82 (1.56 to 4.09) | 0 | | 2 (10) | 3.77 (-0.45 to 8) | 0 |
| Missing outcome data |  |  |  |  |  |  |  |  |  |  |  |  | |  |  |  |
| low risk of bias for missing outcome data | 16 (165) | Random: 1.15 (0.92 to 1.45) Fixed: 1.15 (0.92 to 1.45) | 0 | 13 (88) | Random: 1.16 (0.91 to 1.49) Fixed: 1.16 (0.91 to 1.49) | 0 | 1 (5) | 1.93 (0.83 to 4.48) | 0 | 14 (78) | Random: 2.40 (1.05 to 3.76) Fixed: 2.33 (0.79 to 3.86) | 14 | | 4 (18) | Random: 2.64 (-0.90 to 6.18) Fixed: -0.90 to 6.18) | 0 |
| high risk of bias for missing outcome data | 1 (12) | Fixed: 0.45 (0.12 to 1.74) | 0 | 0 (0) | NA | NA | 0 (0) | NA | NA | NA (NA) | NA | NA | | 0 (0) | NA | NA |
| Intensity of care |  |  |  |  |  |  |  |  |  |  |  |  | |  |  |  |
| inpatient | 17 (164) | Random: 1.11 (0.90 to 1.38) Fixed: 1.11 (0.90 to 1.38) | 15 | 13 (83) | Random: 1.31 (1.01 to 1.70) Fixed: 1.31 (1.01 to 1.70) | 0 | 0 (0) | NA | NA | 0 (0) | NA | NA | | 4 (18) | Random: 2.64 (-0.90 to 6.18) Fixed: -0.90 to 6.18) | 0 |
| outpatient | 0 (0) | NA | NA | 0 (0) | NA | NA | 1 (5) | 1.93 (0.83 to 4.48) | 0 | 16 (87) | Random: 0.85 (-0.16 to 1.86) Fixed: 1.67 (-0.02 to 3.36) | 52 | | 0 (0) | NA | NA |
| **Blinding** | | | | | | | | | | | | | | | | |
| All trials | 23 (217) | Random: 1.00 (0.87 to 1.15) Fixed: 1.00 (0.87 to 1.15) | 0 | 16 (101) | Random: 1.03 (0.84 to 1.26) Fixed: 1.03 (0.83 to 1.28) | 1 | 2 (8) | Random: 1.87 (0.95 to 3.67) Fixed: 1.87 (0.95 to 3.67) | 0 | 19 (104) | Random: 0.47 (-0.38 to 1.32) Fixed: 0.31 (-0.76 to 1.37) | 20 | | 4 (25) | Random: 1.02 (-1.30 to 3.35) Fixed: -0.92 (-6 to 4.15) | 71 |
| **Subgroups** |  |  |  |  |  |  |  |  |  |  |  |  | |  |  |  |
| Trial size |  |  |  |  |  |  |  |  |  |  |  |  | |  |  |  |
| < median | 9 (77) | Random: 0.89 (0.55 to 1.45) Fixed: 0.89 (0.55 to 1.45 | 0 | 6 (24) | Random: 1.51 (0.66 to 3.45) Fixed: 1.51 (0.66 to 3.45) | 0 | 0 (0) | NA | NA | 3 (13) | Random: 1.65 (-4.08 to 7.39) Fixed: 1.65 (-4.08 to 7.39) | 0 | | 2 (11) | Random: -1.05 (-6.34 to 4.12) Fixed: -5.20 (-20.04 to 9.64) | 76 |
| ≥ median | 15 (100) | Random: 0.97 (0.84 to 1.12) Fixed: 0.97 (0.84 to 1.13) | 0 | 9 (47) | Random: 0.97 (0.76 to 1.23) Fixed: 0.96 (0.74 to 1.25) | 16 | 1 (4) | 1.70 (0.85 to 3.41) | 0 | 7 (38) | Random: 1.01 (-0.56 to 2.59) Random: 1.01 (-0.56 to 2.59) | 0 | | 0 (0) | NA | NA |
| Allocation concealment |  |  |  |  |  |  |  |  |  |  |  |  | |  |  |  |
| with allocation concealment | 19 (160) | Random: 0.99 (0.86 to 1.14) Fixed: 0.99 (0.86 to 1.15) | 0 | 14 (79) | Random: 1.03 (0.84 to 1.26) | 1 | 2 (7) | Random: 1.64 (0.66 to 4.10) Fixed: 1.74 (0.58 to 5.27) | 9 | 15 (70) | Random: -1.14 (-2.14 to -0.13) Fixed: -1.14 (-2.14 to -0.13) | 0 | | 2 (16) | Random: -1.46 (-4.84 to 1.92) Fixed: -5.02 (-16.75 to 6.70) | 84 |
| without allocation concealment | 1 (14) | 0.70 (0.17 to 2.87) | NA | 0 (0) | NA | NA | 0 (0) | NA | NA | 1 (4) | 2.86 (-3.57 to 9.28) | 0 | | 0 (0) | NA | NA |
| Missing outcome data |  |  |  |  |  |  |  |  |  |  |  |  | |  |  |  |
| low risk of bias for missing outcome data | 21 (186) | Random: 0.99 (0.86 to 1.14) Fixed: 0.99 (0.85 to 1.15) | 0 | 9 (47) | Random: 0.99 (0.80 to 1.22) Fixed: 0.99 (0.78 to 1.25) | 7 | 1 (5) | 1.73 (0.87 to 3.47) | 0 | 18 (96) | Random: 0.47 (-0.45 to 1.40) Fixed: 0.25 (-0.98 to 1.47) | 26 | | 2 (16) | Random: 1.02 (-1.30 to 3.35) Fixed: -0.92 (-6.00 to 4.15) | 71 |
| high risk of bias for missing outcome data | 1 (14) | Fixed: 0.34 (0.05 to 2.6) | NA | 0 (0) | NA | NA | NA (NA) | NA | NA | NA (NA) | NA | NA | | 0 (0) | NA | NA |
| Intensity of care |  |  |  |  |  |  |  |  |  |  |  |  | |  |  |  |
| inpatient | 19 (173) | Random: 1.01 (0.87 to 1.16) Fixed: 1.01 (0.87 to 1.16) | 0 | 14 (89) | Random: 1.14 (0.92 to 1.41) Fixed: 1.16 (0.91 to 1.47) | 0 | NA (NA) | NA | NA | NA (NA) | NA | NA | | 4 (25) | Random: 1.02 (-1.30 to 3.35) Fixed: 0.92 (-6.00 to 4.15) | 71 |
| outpatient | 0 (0) | NA | NA | 0 (0) | NA | NA | 2 (8) | Random: 1.87 (0.95 to 3.67) Fixed: 1.87 (0.95 to 3.67) | 0 | 19 (102) | Random: 0.21 (-0.69 to 1.11) Fixed: 0.20 (-0.77 to 1.16) | 6 | | 0 (0) | NA | NA |
| ROR: Ratio of odds ratios. ROR > 1 suggests that open-label trials or trials without allocation concealment overestimate beneficial effects of interventions compared to double-blind trials or trials with allocation concealment.  DMD: Difference in mean difference. DMD < 0 suggests that opopen-label trials or trials without allocation concealment overestimate beneficial effects of interventions compared to double-blind trials or trials with allocation concealment.  Number of trials and comparisons in subgroups may not add to the total number of trials and comparisons because excluding trials with various characteristics may render other trials and comparisons uninformative (i.e., comparison does not include at minimum one open-label and one double-blind trial or one trial with and one trial without allocation concealment). | | | | | | | | | | | | | | | | |

### Supplement 8 – The adjusted effects of allocation concealment and blinding on trial results

| **Supp table: Adjusted analyses** | | | | | | | | | | | | | | | |
| --- | --- | --- | --- | --- | --- | --- | --- | --- | --- | --- | --- | --- | --- | --- | --- |
|  | **Mortality** | | | **Mechanical ventilation** | | | **Hospitalization** | | | **Duration of hospitalization** | | | **Duration of mechanical ventilation** | | |
|  | **Comparisons (trials)** | **ROR (95% CI)** | **I2 (%)** | **Comparisons (trials)** | **ROR (95% CI)** | **I2 (%)** | **Comparisons (trials)** | **ROR (95% CI)** | **I2 (%)** | **Comparisons (trials)** | **ROR (95% CI)** | **I2 (%)** | **Comparisons (trials)** | **ROR (95% CI)** | **I2 (%)** |
| **Allocation concealment** | | | | | | | | | | | | | | | |
| number of participants | 15 (193) | Fixed: 1.12 (0.88 to 1.41) Random: 12 (0.88 to 1.41) | 0 | 11 (86) | Fixed: 1.24 (0.90 to 1.70) Random: 1.24 (0.90 to 1.70) | 0 | 1 (5) | Fixed: 1.93 (0.82 to 4.64) | 0 | 13 (80) | Fixed: 1.90 (0.86 to 2.94) Random: 1.90 (0.44 to 3.34) | 32.1 | 1 (12) | Fixed: 3.81 (-0.89 to 8.5) | 0 |
| blinding | 15 (178) | Fixed: 1.14 (0.91 to 1.42) Random: 1.14 (0.91 to 1.42) | 9.7 | 11 (86) | Fixed: 1.20 (0.89 to 1.60) Random: 1.20 (0.89 to 1.60) | 0 | 1 (5) | Fixed: 1.45 (0.45 to 4.70 | 0 | 13 (80) | Fixed: 1.68 (0.69 to 2.67) Random: 1.69 (0.44 to 2.93) | 8.9 | 1 (12) | Fixed: 3.78 (-0.45 to 8.0) | 0 |
| missing data | 15 (178) | Fixed: 1.14 (0.91 to 1.43) Random: 1.14 (0.91 to 1.43) | 0 | 11 (86) | Fixed: 1.20 (0.91 to 1.58) Random: 1.20 (0.91 to 1.58) | 0 | 1 (5) | Fixed: 1.93 (0.83 to 4.5) | 0 | 13 (80) | Fixed: 1.92 (0.92 to 2.91) Random: 1.87 (0.34 to 3.40) | 40.7 | 1 (12) | Fixed: 3.81 (-0.39 to 8.1) | 0 |
| inpatient vs. outpatient | 15 (178) | Fixed: 1.25 (0.91 to 1.40) Random: 1.25 (0.91 to 1.40) | 19.5 | 11 (86) | Fixed: 1.20 (0.92 to 1.58) Random: 1.20 (0.92 to 1.58) | 0 | 1 (5) | Fixed: 1.93 (0.83 to 4.5) | 0 | 13 (80) | Fixed: 1.23 (0.36 to 2.10) Random: 1.43 (-0.15 to 3.02) | 59.3 | 1 (12) | Fixed: 3.81 (-0.39 to 8.1) | 0 |
| all of the above | 11 (151) | Fixed: 1.15 (0.88 to 1.51) Random: 1.15 (0.88 to 1.51) | 0 | 8 (70) | Fixed: 1.00 (0.97 to 1.04) Random: 1.00 (0.97 to 1.04) | 0 | 1 (5) | Fixed: 1.43 (0.44 to 4.63) | 0 | 6 (50) | Fixed: 2.84(0.57 to 5.11) Random: 2.25 (-1.56 to 6.1) | 39.7 | 1 (12) | Fixed: 3.5 (-1.74 to 8.84) | 0 |
| **Blinding** | | | | | | | | | | | | | | | |
| number of participants | 19 (200) | Fixed: 1.00 (0.85 to 1.18) Random: 1.00 (0.85 to 1.18) | 0 | 12 (89) | Fixed: 1.11 (0.85 to 1.44) Random: 1.11 (0.85 to 1.44) | 0 | 1 (5) | Fixed: 1.85 (0.86 to 3.96) | 0 | 15 (92) | Fixed: 1.75 (0.02 to 3.50) Random: 1.72 (-0.12 3.60) | 31.6 | 2 (19) | Fixed: -0.06 (-4.01 to 3.89) Random: -3.43 (-15.6 to 8.7) | 81.4 |
| blinding | 19 (216) | Fixed: 0.98 (0.85 to 1.13) Random: 0.98 (0.85 to 1.13) | 0 | 12 (89) | Fixed: 1.11 (0.85 to 1.44) Random: 1.11 (0.85 to 1.44) | 0 | 1 (5) | Fixed: 1.85 (0.86 to 3.96) | 0 | 15 (92) | Fixed: 0.42 (-0.57 to 1.40) Random: 0.42 (-0.63 to 1.48) | 0 | 2 (19) | Fixed: -1.46 (-4.84 to 1.91) Random: -16.75 to 6.7) | 83.6 |
| missing data | 19 (216) | Fixed: 0.98 (0.85 to 1.13) Random: 0.98 (0.84 to 1.14) | 0 | 12 (89) | Fixed: 1.11 (0.85 to 1.44) Random: 1.11 (0.85 to 1.44) | 0 | 1 (5) | Fixed: 1.73 (0.87 to 3.5) | 0 | 15 (92) | Fixed: 0.95 (-0.06 to 1.95) Random: 0.95 (-0.06 to 1.95) | 0 | 2 (19) | Fixed: -1.15 (-4.5 to 2.2) Random: -4.9 (-16.9 to 7.20) | 84.5 |
| inpatient vs. outpatient | 19 (216) | Fixed: 0.99 (0.87 to 1.15) Random: 0.99 (0.87 to 1.15) | 0 | 12 (89) | Fixed: 1.04 (0.84 to 1.30) Random: 1.04 (0.83 to 1.30) | 0 | 1 (5) | Fixed: 1.73 (0.87 to 3.5) | 0 | 15 (92) | Fixed: -0.60 (-0.44 to 1.63) Random: -0.60 (-0.44 to 1.63) | 0 | 2 (19) | Fixed: -1.15 (-4.5 to 2.2) Random: -4.9 (-16.9 to 7.20) | 84.5 |
| all of the above | 16 (181) | Fixed: 0.90 (0.75 to 1.07) Random: 0.89 (0.75 to 1.07) | 0 | 10 (80) | Fixed: 1.02 (0.75 to 1.4) Random: 1.02 (0.75 to 1.4) | 0 | 1 (5) | 1.50 (0.53 to 4.20) | 0 | 8 (62) | Fixed: -0.52 (-1.82 to 2.86) Random: 0.38 (-2.53 to 3.30) | 0 | 2 (19) | Fixed: -1.72 (-6.07 to 2.6) Random: -4.14 (-14.8 to 6.5) | 74.8 |

### Supplement 9 – The effects of allocation concealment and blinding on trial results excluding trials with unclear reporting of allocation concealment or blinding status

| **Outcomes** | **Comparisons (trials)** | **ROR or DMD (95% CI)** | **I2 (%)** |
| --- | --- | --- | --- |
| **Allocation** | | | |
| **Mortality** | 3 (30) | Random: 0.77 (0.17 to 3.55) Fixed: 0.76 (0.32 to 1.82) | 64.3 |
| **Mechanical ventilation** | 1 (7) | Fixed: 0.32 (0.05 to 2.22) | NA |
| **Hospitalization** | 0 (0) | NA | NA |
| **Duration of hospitalization** | 1 (8) | 3.99 (-15.21 to 23.20) | NA |
| **Duration of mechanical ventilation** | 1 (3) | -1.34 (-11.00; 8.32) | NA |
| **Blinding** | | | |
| **Mortality** | 38 (230) | Random: 1.03 (0.91 to 1.15) Fixed: 1.03 (0.91 to 1.15) | 0 |
| **Mechanical ventilation** | 16 (100) | Random: 1.03 (0.83 to 1.28) Fixed: 1.03 (0.84 to 1.26) | 0.7 |
| **Hospitalization** | 2 (8) | Random: 1.87 (0.95 to 3.67) Fixed: 1.87 (0.95 to 3.67) | 0 |
| **Duration of hospitalization** | 19 (104) | Random: 0.31 (-0.76 to 1.37) Fixed: 0.47 (-0.38 to 1.32) | 20.4 |
| **Duration of mechanical ventilation** | 4 (25) | Random: -0.92 (-6.00 to 4.15) Fixed: 1.02 (-1.30 to 3.35) | 71.2 |

### Supplement 10– Estimated ratios of odds ratios (RORs) and differences in mean differences (DMDs) comparing trials in which patients and healthcare providers are blinded and open-label trials

#### Mechanical ventilation

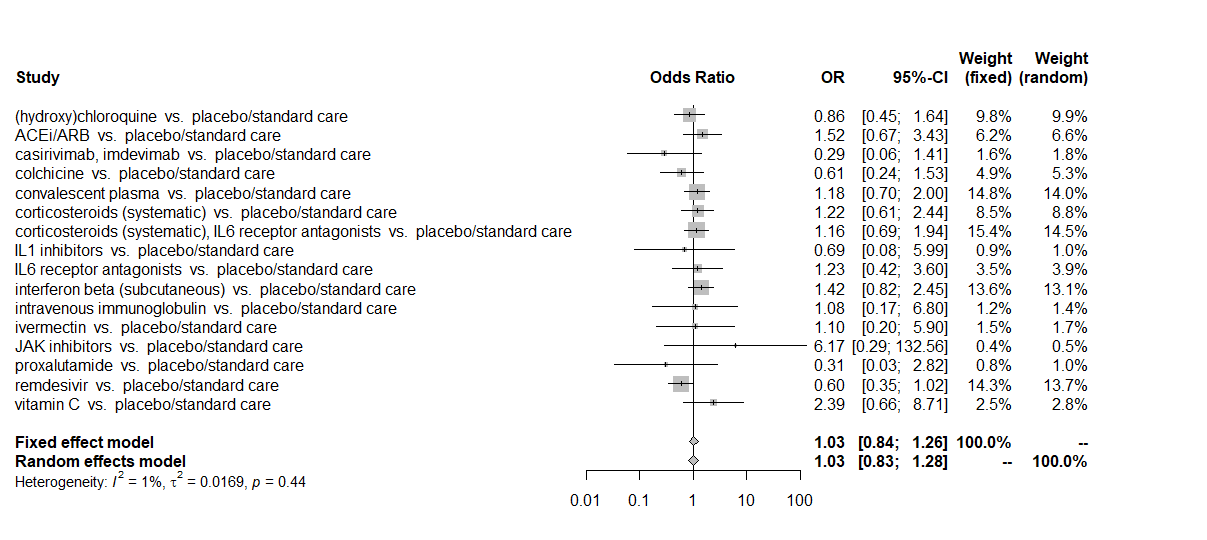

#### Hospitalization

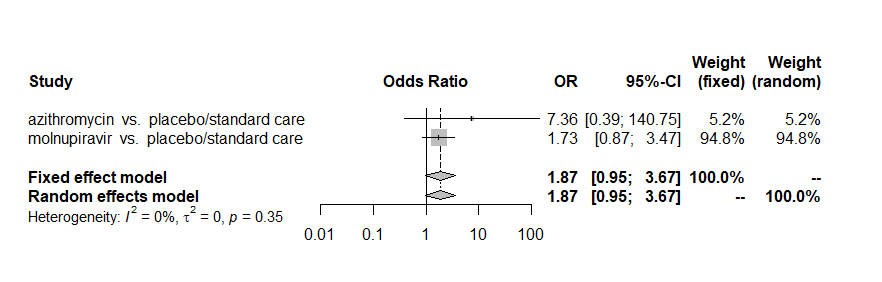

#### Duration of hospitalization

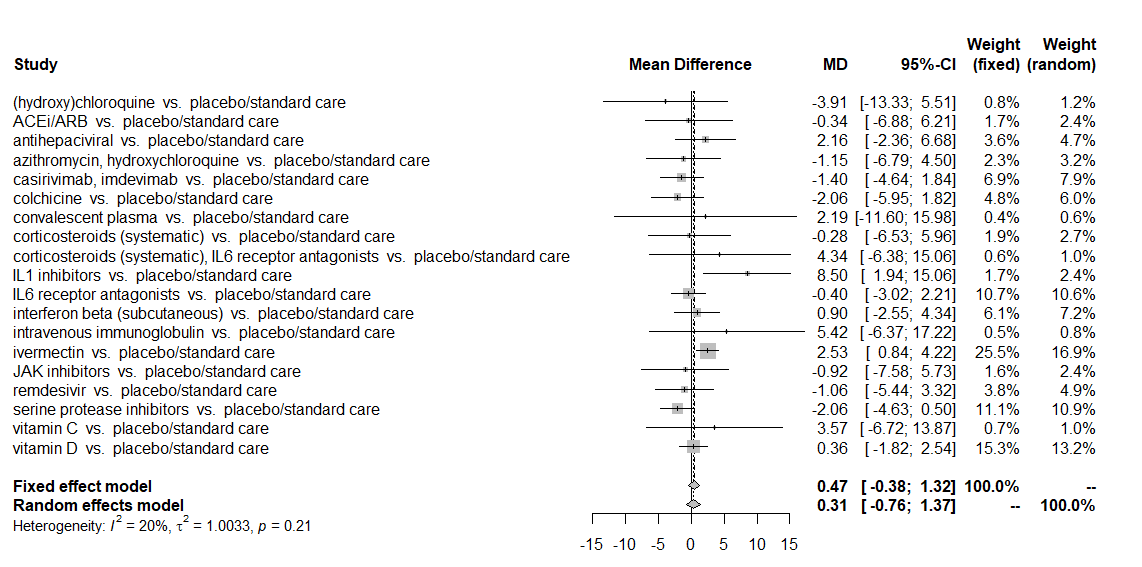

#### Duration of mechanical ventilation

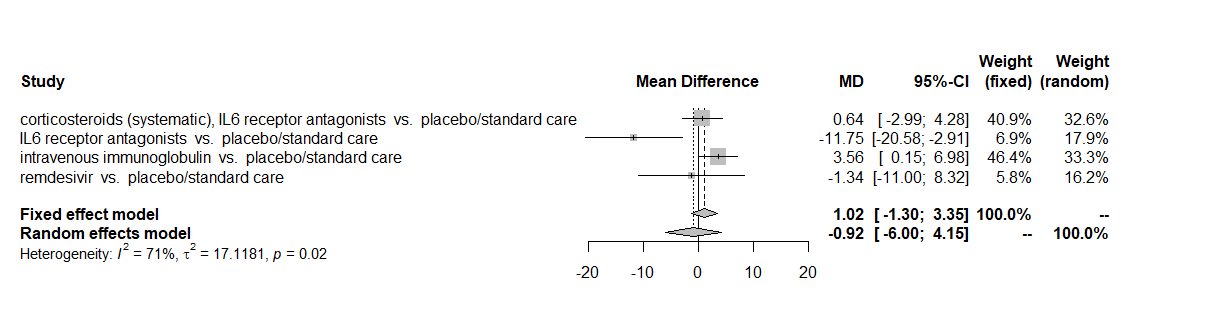
